# Genome-wide rare variant score associates with morphological subtypes of autism spectrum disorder

**DOI:** 10.1101/2021.10.20.21264950

**Authors:** Ada J.S. Chan, Worrawat Engchuan, Miriam S. Reuter, Zhuozhi Wang, Bhooma Thiruvahindrapuram, Brett Trost, Thomas Nalpathamkalam, Carol Negrijn, Sylvia Lamoureux, Giovanna Pellecchia, Rohan Patel, Wilson W.L. Sung, Jeffrey R. MacDonald, Jennifer L. Howe, Jacob Vorstman, Neal Sondheimer, Nicole Takahashi, Judith H. Miles, Evdokia Anagnostou, Kristiina Tammimies, Mehdi Zarrei, Daniele Merico, Dimitri J. Stavropoulos, Ryan K.C. Yuen, Bridget A. Fernandez, Stephen W. Scherer

**Affiliations:** The Centre for Applied Genomics, Genetics and Genome Biology, The Hospital for Sick Children, Toronto, ON, Canada; Genetics and Genome Biology, The Hospital for Sick Children, Toronto, ON, Canada; Ted Rogers Centre for Heart Research, The Hospital for Sick Children, Toronto, ON, Canada; Provincial Medical Genetics Program, Eastern Health, St. John’s, NL, Canada; Department of Psychiatry, The Hospital for Sick Children, Toronto, ON, Canada; Department of Psychiatry, University of Toronto, Toronto, ON, Canada; Department of Molecular Genetics, University of Toronto, Toronto, ON, Canada; Division of Clinical and Metabolic Genetics, Department of Pediatrics, The Hospital for Sick Children, Toronto, ON, Canada; Department of Pediatrics, University of Toronto, Toronto, ON, Canada; Thompson Center for Autism and Neurodevelopmental Disorders, University of Missouri, Columbia, MO, USA; Holland Bloorview Kids Rehabilitation Hospital; Department of Women’s and Children’s Health, Karolinska Institutet, Stockholm, Sweden; Deep Genomics Inc., Toronto, ON, Canada; Department of Paediatric Laboratory Medicine, Genome Diagnostics, The Hospital for Sick Children, Toronto, ON Canada; Department of Paediatric Laboratory Medicine, Genome Diagnostics, Hospital for Sick Children, Toronto, Ontario, Canada; Department of Pediatrics and The Saban Research Institute, Children’s Hospital Los Angeles, Keck School of Medicine of University of Southern California, Los Angeles CA USA; Discipline of Genetics, Faculty of Medicine, Memorial University of Newfoundland, St. John’s NL Canada; McLaughin Centre, University of Toronto, Toronto, ON, Canada

## Abstract

Defining different genetic subtypes of autism spectrum disorder (ASD) can enable the prediction of developmental outcomes. Based on minor physical and major congenital anomalies, we categorized 325 Canadian children with ASD into dysmorphic and nondysmorphic subgroups. We developed a method for calculating a patient-level, genome-wide rare variant score (GRVS) from whole-genome sequencing (WGS) data. GRVS is a sum of the number of variants in morphology-associated coding and non-coding regions, weighted by their effect sizes. Probands with dysmorphic ASD had a significantly higher GRVS compared to those with nondysmorphic ASD (*P*= 0.027). Using the polygenic transmission disequilibrium test, we observed an over-transmission of ASD-associated common variants in nondysmorphic ASD probands (*P*= 2.9×10^−3^). These findings replicated using WGS data from 442 ASD probands with accompanying morphology data from the Simons Simplex Collection. Our results provide support for an alternative genomic classification of ASD subgroups using morphology data, which may inform intervention protocols.

## Main

Autism spectrum disorder (ASD), which is diagnosed on the basis of behavioral assessments that reveal social communication deficits and repetitive behaviors, is often associated with traits including major congenital anomalies (MCAs), minor physical anomalies (MPAs)^1,2^ and intellectual disability^3-5^. Increasingly, penetrant variants of diagnostic value^6,7^ and lesser impact common variants are being implicated in the etiology of ASD^4,8^.

Autistic individuals who are more dysmorphic (complex ASD) tend to have lower intelligence quotients (IQ) and more brain and other major congenital anomalies^9,10^ compared with those who are less dysmorphic (essential ASD), leading to poorer developmental outcomes. Individuals with complex ASD are also less likely to have a family history of ASD, suggesting that morphological subtypes can reveal informative genetic differences among ASD subgroups^9^.

Genetic liability to ASD can be quantified using a polygenic risk score (PRS), which is a weighted sum of ASD-associated common variants, using effect sizes drawn from genome-wide association studies^11^. A similar score for rare variants remains to be established. Rare variant studies use burden analyses to compare the frequency of rare variants, equally weighted, between cases and controls or among ASD subtypes^3,12,13^. Quadratic tests have also been used in rare variant association tests and typically weigh variants by minor allele frequency^14,15^. However, effect sizes depend on the affected gene and variant type, and these variables should be considered in rare variant analyses.

Here, from two independent cohorts, we used whole-genome sequences (WGS) and detailed clinical morphology data to: 1) develop a genome-wide rare variant score (GRVS) to measure the relationship between rare variants and morphology, and 2) examine the contribution of rare and common variants in morphological ASD subtypes (Figure 1 and Supplementary figure 1).

**Figure 1:**
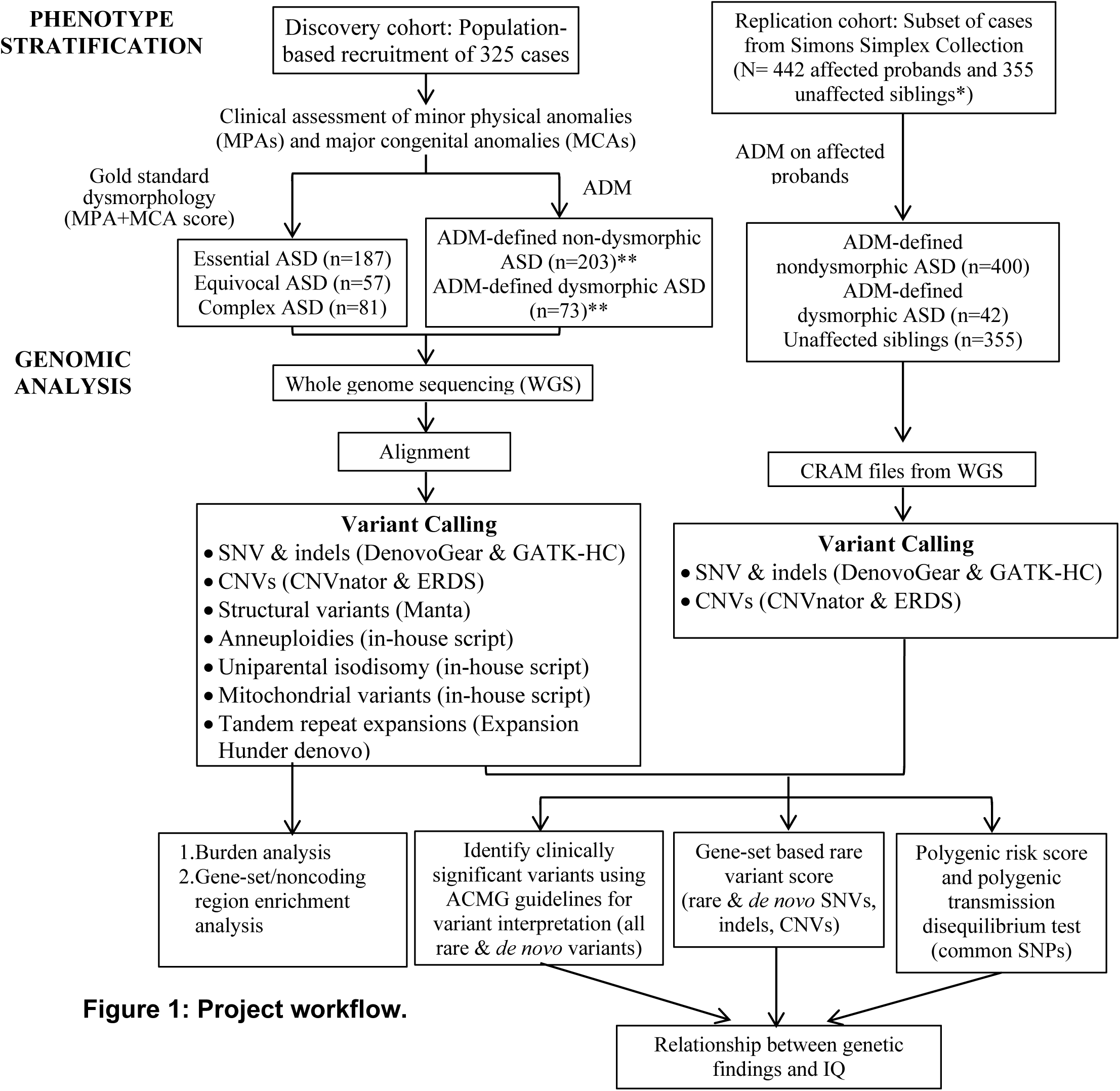
Project workflow. Summary of phenotype stratification, whole-genome sequencing workflow, and genomic analyses performed in this study. ASD, Autism spectrum disorder; ADM, Autism dysmorphology measure; CNVs, copy number variants; SNVs, single nucleotide variants; indels, insertions and deletions; ERDS, estimation by read depth with single nucleotide variants; GATK-HC, Genome Analysis Toolkit-Haplotype Caller; SNPs, single nucleotide polymorphisms; ACMG, American College of Medical Genetics and Genomics; IQ, Intelligence quotient. *Unaffected siblings were used for GRVS and PRS analyses. **Excluding samples with false negative ADM-defined nondysmorphic ASD. We also included only samples sequenced on Illumina platforms to be consistent with replication cohort. For variant calling, on average per sample, we detected ∼3.7 million SNPs, 36,514 rare single nucleotide variants (SNVs), 4,113 small insertions and deletions (indels), 13 rare copy number variants (CNVs), 390 rare SVs, 73.4 *de novo* SNVs, 7.3 *de novo* indels, and 0.1 *de novo* CNVs (Supplementary Table 2). Experimental validation rates were 94.8%, 85.7%, and 87.5%, respectively, for *de novo* SNVs, indels, and CNVs (Supplementary Tables 3 and 4). Using GRVS, we were able to quantify and validate the contribution of morphology-associated, rare sequence-level and copy number variants to morphological ASD subtypes. While we can call other SVs from the WGS, there needs to be higher-quality data before these can be effectively incorporated into GRVS.

For our discovery cohort, we used a population-based sample of 325 unrelated children with Autism Diagnostic Observation Scale (ADOS)-confirmed ASD. Following clinical examination, a total morphology score was assigned to each case based on the number of MPAs and MCAs^9,10^. The cohort was then stratified into three subtypes of increasing morphologic severity: 187 essential ASD (57.5%), 57 equivocal ASD (17.5%) and 81 complex ASD (24.9%) (Supplementary Table 1). We further stratified these samples into two subtypes by combining complex and equivocal ASD into a single dysmorphic ASD grouping and redefining essential ASD as nondysmorphic ASD.

We performed WGS on 795 genomes (325 probands and 470 parents) and detected all classes of variation (SNV, indel, CNV and structural variants (SVs) (Figure 1, Supplementary Table 2-4). Using the American College of Medical Genetics and Genomics guidelines^16,17^, we identified a total of 46 clinically significant variants (CSVs) in 46 of 325 probands (14.1%) (Supplementary Tables 5-7). The proportion of dysmorphic ASD cases with a CSV (25.9%; 35/135) was significantly higher than nondysmorphic ASD (5.8%, 11/190) (*P*= 3.2×10^−7^, one-sided Fisher’s test), consistent with our previous findings^10^. We also identified 29 variants of uncertain significance (VUS) in 26 probands that were of interest, including tandem repeat expansions in previously reported ASD candidate loci^18^; three probands each had two VUS (Supplementary Tables 5-7 and Supplementary Note).

To further investigate the contribution of rare variants among morphological ASD subtypes, we first conducted a rare variant burden analysis and multiple test correction using the Benjamini Hochberg approach (BH-FDR) (see methods). We found a significantly higher prevalence of rare coding deletions >10kb in probands with more dysmorphic features (*P=* 5.00 × 10^−4^ and BH-FDR= 5.00 × 10^−3^, Supplementary Table 8). Rare coding duplications >10kb and ≤10kb, genic deletions ≤10kb, loss-of-function (LoF) and missense variants were not significantly different among subtypes (Supplementary Table 8).

We then performed enrichment and burden analyses to identify gene sets or noncoding regions, respectively, that were differentially affected by rare or *de novo* variants between the morphological ASD subtypes. The 67 gene sets and noncoding regions studied have been previously associated with ASD^13,19-23^. After multiple-testing correction (permutation-based false discovery rate (FDR) <20%), 20 significant gene sets or noncoding regions were identified (Supplementary Tables 9 and 10). We observed that probands with dysmorphic features had higher burdens of deletions and missense variants impacting genes responsible for various neuronal functions and duplications >10kb impacting brain-expressed genes (Figure 2 and Supplementary Table 9). Dysmorphic probands also had a significantly higher prevalence of rare deletions ≤10kb overlapping promoters of long noncoding genes and duplications (larger and smaller than 10kb) overlapping active brain enhancers (Supplementary Figure 2, Supplementary Table 10).

**Figure 2:**
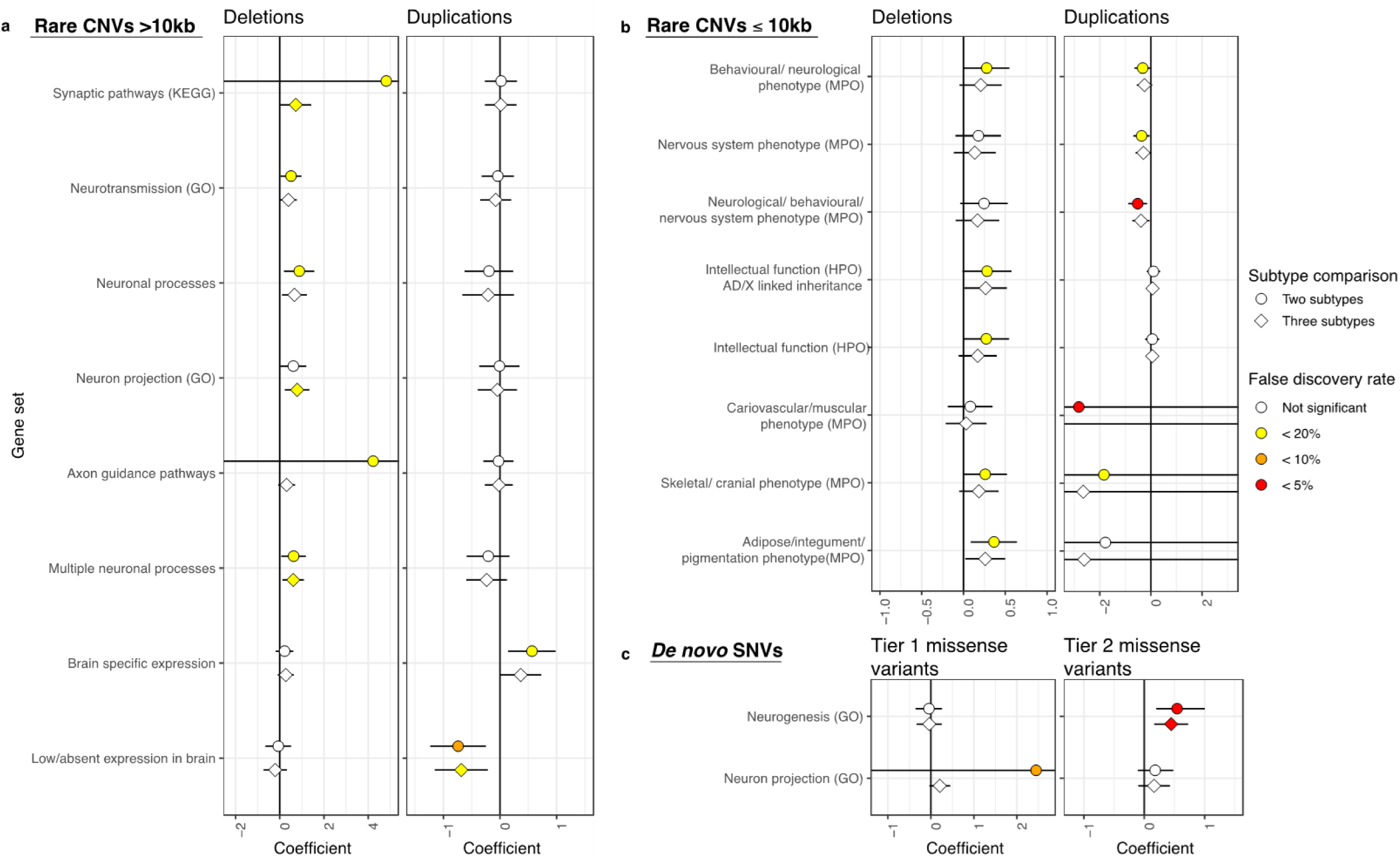
Gene sets for which *de novo* and rare coding variants are significantly more prevalent in some subtypes of ASD. We define events as (**a**,**b**) genes impacted by CNVs or (**c**) as variants for SNVs and indels. The coefficient is the relationship between the number of events in each gene set and the ASD subtypes; it reflects the effect size of a variant type and gene set among different ASD subtypes. Positive coefficients indicate more events in individuals with ASD and more dysmorphic features; negative coefficients indicate more events in individuals with ASD and fewer dysmorphic features. We show only gene sets for which **a**,**b**) rare CNVs, or **c**) *de novo* missense variants are significantly more prevalent in different subtypes of ASD. Tier 1 and 2 missense variants consist of all or only predicted damaging missense variants, respectively, as defined in Yuen, *et al*.^23^ Symbol shapes indicate the subtype comparisons that were conducted for each combination of gene set and variant type. Two subtype comparison= nondysmorphic vs. dysmorphic ASD. Three subtype comparison= essential vs. equivocal vs. complex ASD. Coloured shapes indicate significant signals after multiple test correction by permutation-based FDR. Error bars indicate 95% confidence intervals.

We then tested the collective contribution of rare variants in morphology-associated regions, while considering the effect size of each variant, which varies depending on the variant type and morphology-associated region. We developed a GRVS for each proband, which is a weighted sum of the number of rare variants in morphology-associated regions identified from gene set enrichment and noncoding burden tests (Supplementary Table 11). We weighed the number of rare variants in each morphology-associated region as well as the variant type (i.e., coding or noncoding deletions and duplications >10kb or ≤10kb, loss-of-function variants, missense variants, and noncoding SNVs and indels) using the coefficients from logistic regression models.

To calculate GRVSs for each proband in the discovery cohort, we used a 10-fold cross validation strategy to reduce over-fitting (Supplementary Figure 1a). We used Nagelkerke’s R^2^ to determine the optimal *P* value threshold (*P*<0.1) to identify morphology-associated regions (Supplementary Figure 3a and Online Methods). GRVS can be calculated for probands regardless of whether their parents have been sequenced. However, there would be a systematic difference in GRVSs in this cohort if all probands were used because those probands whose parents have been sequenced would include scores from *de novo* variants, whereas those without sequenced parents would not have *de novo* variant scores. To avoid this, GRVS was calculated only for probands with two sequenced parents (n= 235) (Figure 3a and Supplementary Table 12).

**Figure 3:**
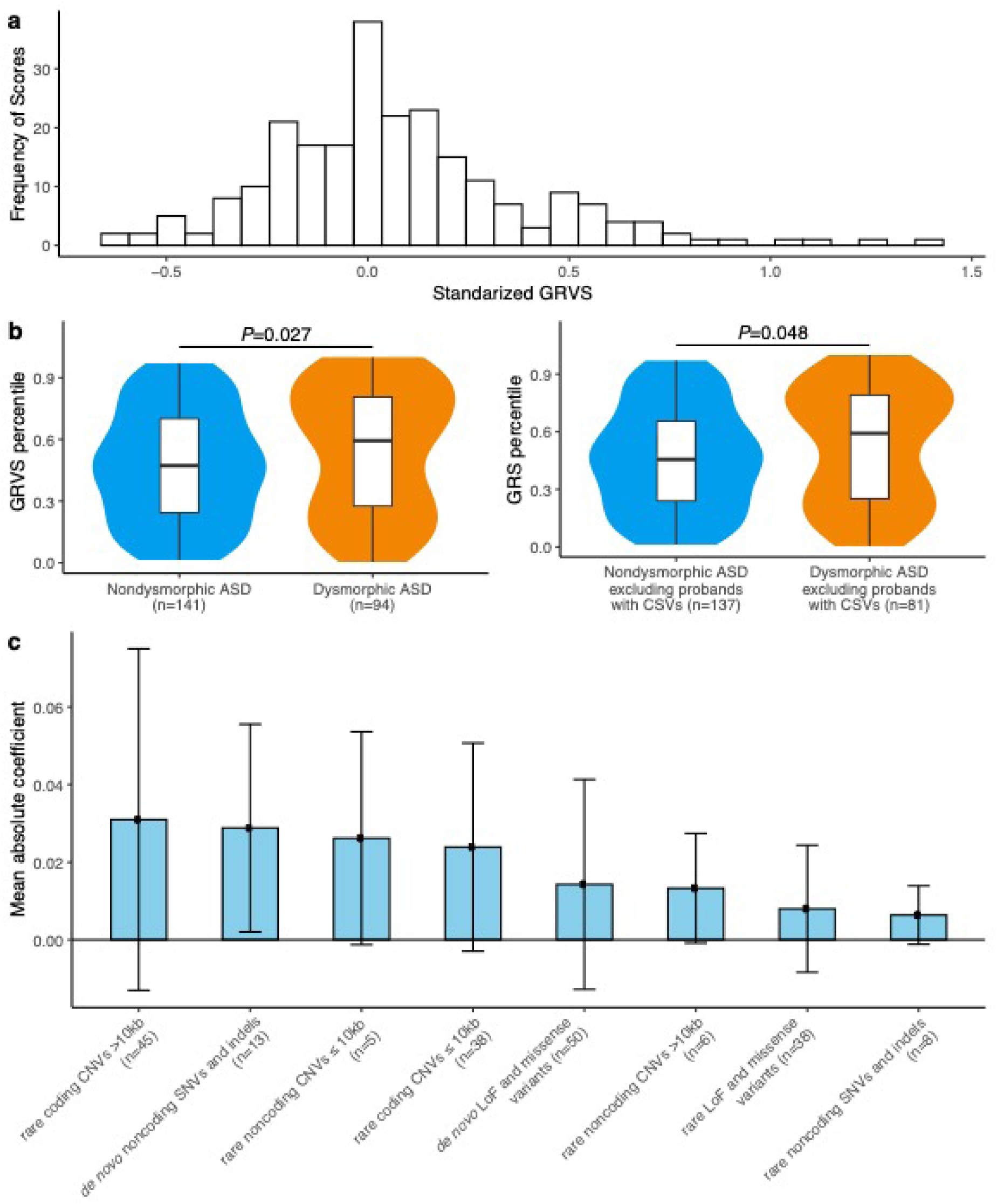
Genome-wide rare variant score in ASD subtypes. Events are comprised of variants for SNVs and indels or genes impacted by CNVs. For each sample, the GRVS is the sum of rare and *de novo* events in morphology-associated regions, weighted by effect size (estimated from the coefficients in the regression model). GRVSs were generated 30 times for each sample (see methods), yielding an average score and average number of variants. CSVs, clinically significant variants. **a**) Distribution of standardized GRVS for the discovery cohort (n=235). **b**) GRVSs for the whole cohort (left plot, n=235) or the whole cohort excluding the 17 probands with clinically significant variants (right plot, n=218), were ordered and ranked by percentile. Note that while 46 probands in the discovery cohort (n=325) had CVSs, only 17 of them had two sequenced parents meeting inclusion criterion for the GRVS group (n=235). Violin plots show the distributions of the samples’ GRVS percentiles; box plots contained within show the median and quartiles of the percentiles for each subtype. *P* values denote the probability that the GRVS in dysmorphic ASD is not greater than nondysmorphic ASD (one-sided, Wilcoxon rank sum test). **c**) Rare variants have different effect sizes. The mean coefficient reflects the effect size of a variant type. Coefficients of deletions and duplications of the same size bin were averaged together. Coefficients of predicted LoF variants, missense variants, and predicted damaging missense variants were averaged together. Error bars indicate mean ± standard deviation. The number of morphology-associated regions for each variant type is indicated the y-axis with “n= ”.

Probands with dysmorphic ASD had significantly higher average GRVSs than those with nondysmorphic ASD (*P=* 0.027, one-sided Wilcoxon rank sum test) (Figure 3b). Most probands (95.7%, 225/235) had more than one variant impacting morphology-associated regions (Supplementary Table 12). Rare coding CNVs had the highest effect size; rare noncoding SNVs and indels had the lowest (Figure 3c and Supplementary Table 11).

Using the GRVS formula, we calculated a score for CSVs that overlapped an ASD relevant, morphology-associated region (so that effect size was available for calculation) and that occurred in probands with sequencing data from both parents. 17 of the 46 CSVs met these criteria. No score was calculated for the remaining 29 variants because 15 were identified in probands where both parents were not available for sequencing, and 14 variants were not located in or encompassed by one of the 20 morphology-associated regions. (Online Methods). In 47% of samples with CSV scores (8/17 probands, Supplementary Table 13), CSVs contributed >50% of the total GRVS. When we excluded the probands with CSVs, those with dysmorphic ASD still had significantly higher average GRVSs than those with nondysmorphic ASD (*P=* 0.048, one-sided Wilcoxon rank sum test, Figure 3b). These findings suggest that variants in morphology-associated regions that are not CSVs also significantly contribute to morphological outcomes in ASD.

To explore the contribution of common (minor allele frequency >0.05) ASD-associated variants in ASD subtypes, we calculated polygenic risk scores (PRS) for ASD and body mass index (BMI)^8^ (Online Methods and Supplementary Table 12). We then compared these scores across the morphologic groups using the polygenic transmission disequilibrium test (pTDT)^8^, which compares the PRS of the proband to parents’ mean PRS. We found a significant over-transmission of common ASD-associated variants in probands with nondysmorphic ASD (*P*= 2.9×10^−3^, one-sided t-test) and no significant over-transmission in probands with dysmorphic ASD (*P=* 0.3) (Figure 4). PRS for BMI was selected as a negative control because there is no genetic correlation between BMI and ASD^24^, and we did not find over-transmission of PRS for BMI in either subtype (Figure 4).

**Figure 4:**
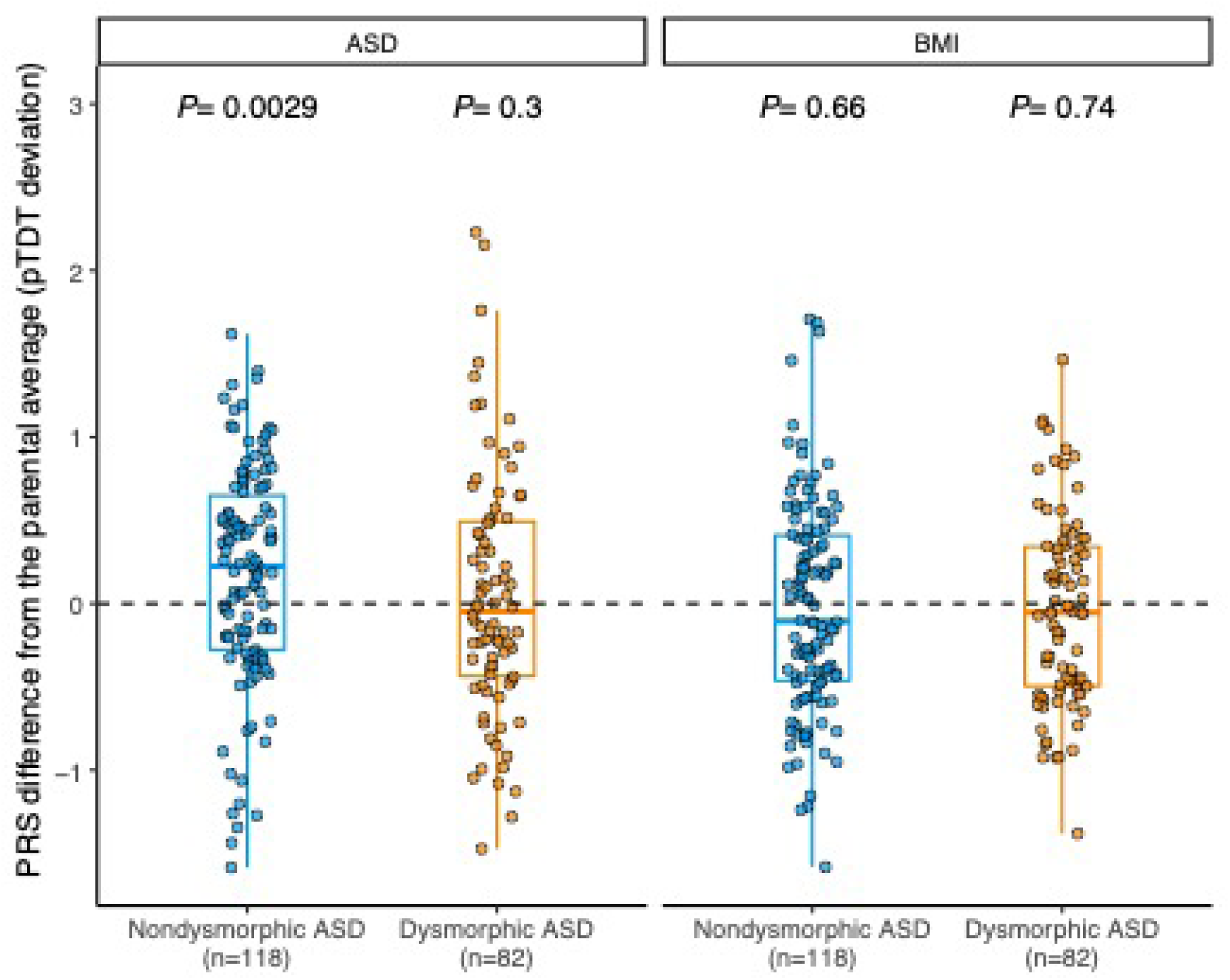
Inheritance of polygenic risk for ASD and BMI in morphologic subtypes. Differences in polygenic risk score (PRS) for ASD and BMI between subjects and their respective mid-parent score. Box plots depict the median and quartiles of polygenic transmission disequilibrium test (pTDT) deviation. Dots represent pTDT deviations of subjects. *P* values for each subgroup indicate the probability that the mean of the pTDT deviation distribution is not greater than zero (one-sided, *t*-test), as depicted by the dotted line.

IQ is often negatively correlated with the burden of rare variants^3,4,13,25,26^. We therefore examined our probands with dysmorphic ASD and determined they had a significantly lower mean IQ compared to nondysmorphic ASD (*P=* 0.013, one-sided t-test, Figure 5a and Supplementary Table 12). Probands with a CSV had significantly lower IQ compared to probands without a CSV (*P=* 2.2 ×10^−4^, one-sided t-test, Figure 5b). However, IQ was not significantly correlated with GRVS (rho= -0.042, *P=* 0.64, Figure 5c) or PRS (rho= -0.15, *P=* 0.12, Figure 5d).

**Figure 5:**
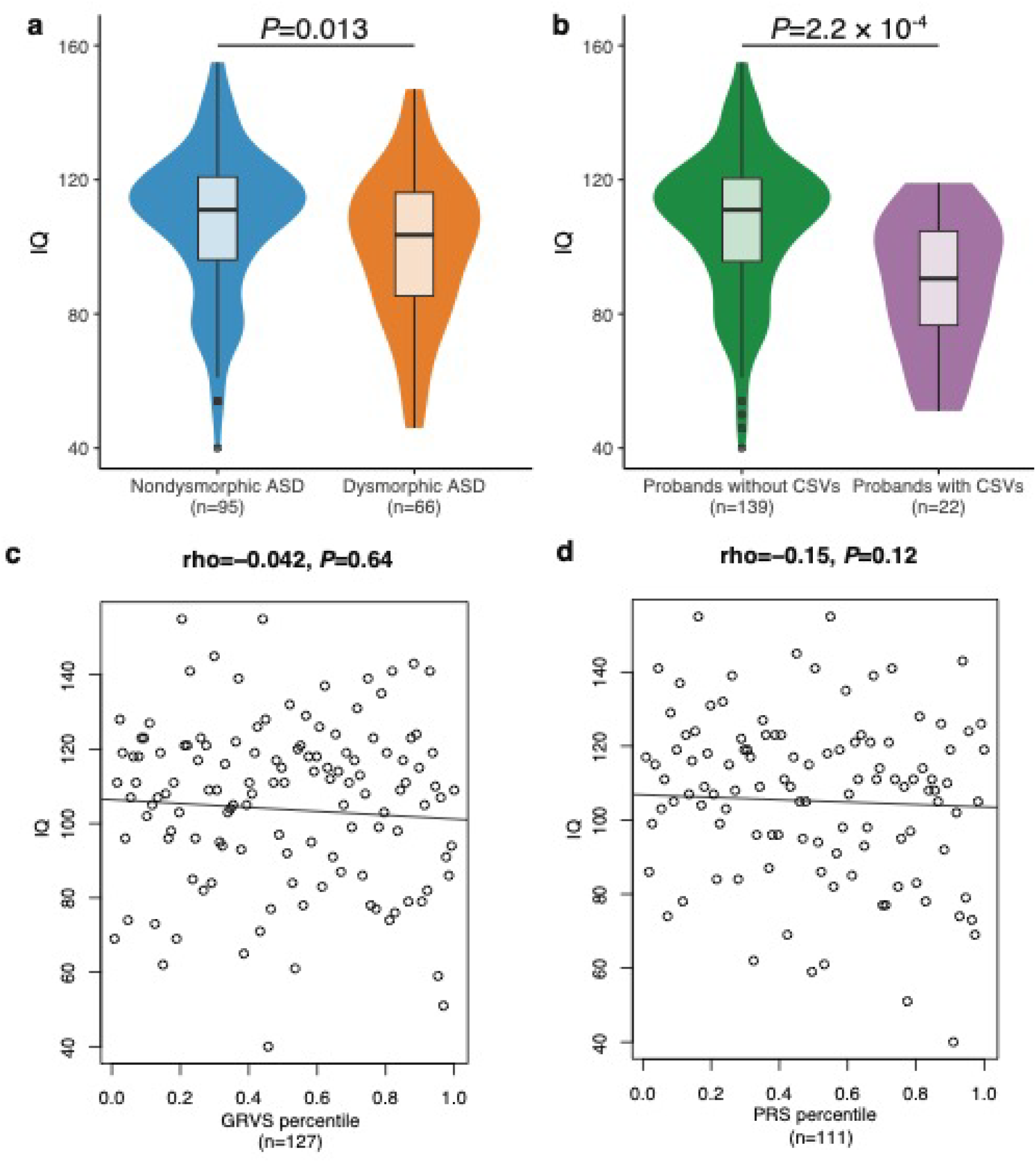
Relationship between IQ, morphological ASD subtypes and genetic variants. a) Comparison of IQ among morphological ASD subtypes. b) Comparison of IQ between probands with and without a CSV. a,b) Violin plots show the distributions of the probands’ IQ; box plots contained within show the median and quartiles of IQ for each subtype. *P* values denote the probability that the mean IQ of nondysmorphic ASD or probands without CSVs is not greater than dysmorphic ASD or probands with CSVs, respectively (one-sided, t-test). Correlation between IQ and c) GRVS and d) PRS is shown. c,d) Each dot represents the IQ and GRVS or PRS percentile of a sample. The linear regression line indicates the linear correlation between IQ and GRVS or PRS percentiles. Correlation coefficient is quantified by Spearman’s rho correlation. *P* values indicate the probability that the correlation is occurred due to chance.

We repeated our analysis on a replication cohort of relevant samples from the Simons Simplex Collection (442 ADOS-confirmed affected probands and 355 unaffected siblings)^27^. The affected probands had been categorized into two morphological subtypes (400 nondysmorphic and 42 dysmorphic cases)^27^ using the Autism Dysmorphology Measure (ADM)^28^. In contrast to the discovery cohort, the SSC probands were classified by targeted physical examinations performed by individuals without expert training in dysmorphogy, and the classification did not incorporate the presence or absence of major congenital anomalies. To compare the two cohorts, we reclassified a subset of the original discovery cohort based on minor anomalies alone using the ADM algorithm (203 nondysmorphic and 73 dysmorphic cases, Online Methods). We calculated new GRVSs for the ADM-reclassified discovery cohort using a 10-fold cross-validation approach (143 nondysmorphic and 48 dysmorphic cases met criteria for inclusion in this analysis, Supplementary Figure 1a, Supplementary Tables 12 and 14, and Online Methods). We used Nagelkerke’s R^2^ to determine the optimal *P*-value threshold and identified 32 morphology-associated regions, which largely overlapped with our original analysis (Supplementary Figure 3b). The morphology-associated regions (*P*< 0.1, Supplementary Table 15) identified in the reclassified discovery cohort were used to calculate GRVSs for the replication cohort (Supplementary Figure 1b, Supplementary Table 16, and Online Methods).

In both cohorts, probands with ADM-defined dysmorphic ASD had significantly higher GRVSs (*P*_discovery=_ 3.6 ×10^−6^ and *P*_replication=_ 2.7 ×10^−4^, one-sided Wilcoxon rank sum test, Figure 6a) and yield of CSVs (*P*_discovery=_ 2.7 ×10^−7^ and *P*_replication=_ 2.1 ×10^−3^, one-sided Wilcoxon rank sum test, Figure 6b and Supplementary Tables 17 and 18) compared to ADM-defined nondysmorphic ASD, consistent with our findings using the gold-standard dysmorphology classification. In the replication cohort, unaffected siblings had a significantly lower GRVS compared to ADM-defined dysmorphic ASD (*P=* 7.7 ×10^−4^ one-sided Wilcoxon rank sum test) but did not have a significantly lower GRVS compared to ADM-defined nondysmorphic ASD (Figure 6a). Furthermore, unaffected siblings of nondysmorphic probands did not have a significantly lower GRVS compared unaffected siblings of dysmorphic probands (*P=* 0.75, one-sided Wilcoxon rank sum test, data not shown).

**Figure 6:**
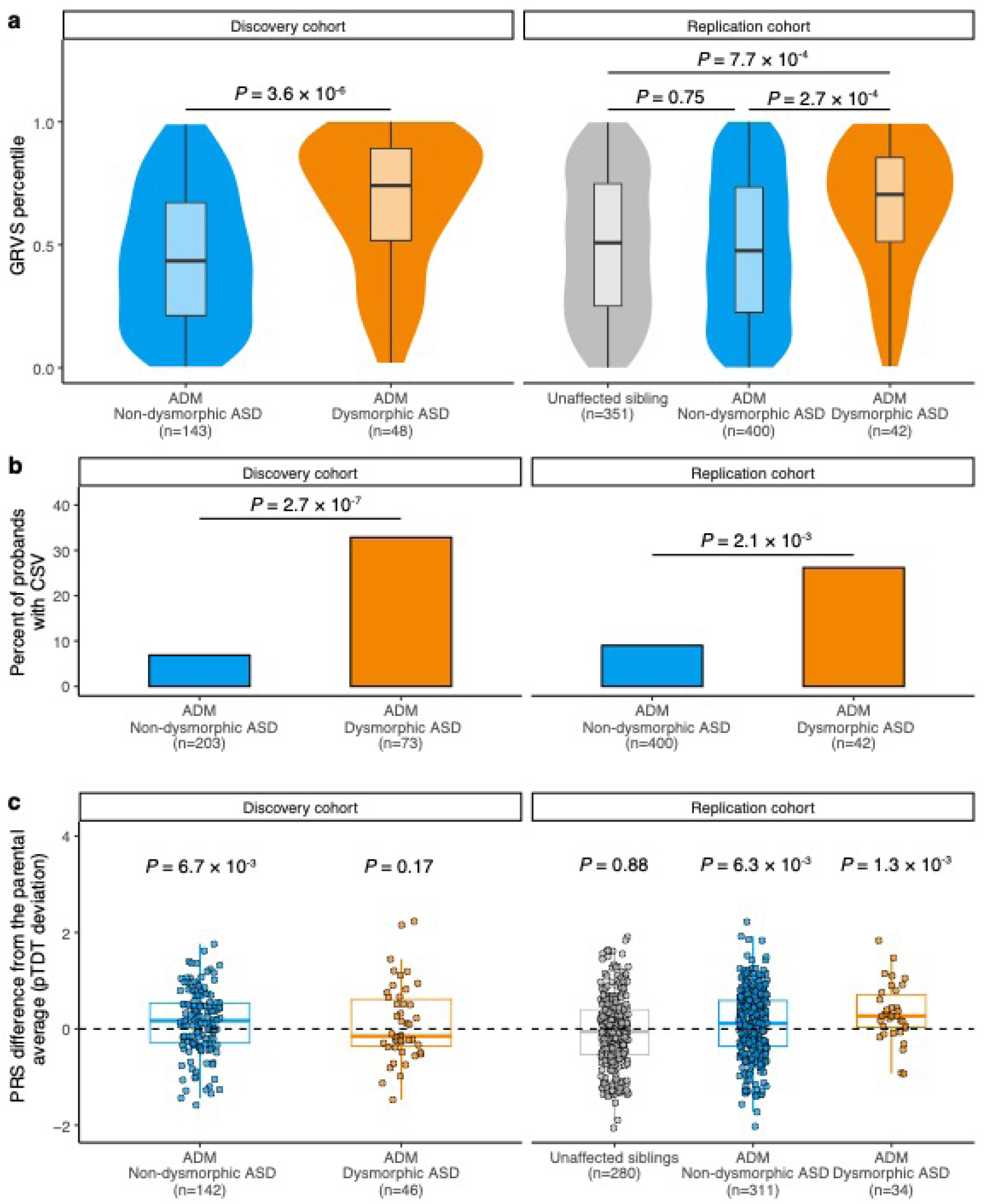
Replication of rare and common genetic findings in subset of Simons Simplex Collection cohort. a) GRVSs for each cohort were ordered and ranked by percentile. Violin plots show the distributions of the probands’ GRVS percentiles; box plots contained within show the median and quartiles of the percentiles for each subtype. *P* values denote the probability that the GRVS in ADM-defined dysmorphic ASD is not greater than ADM-defined nondysmorphic ASD (one-sided, Wilcoxon rank sum test). b) Yield of CSVs between dysmorphic and nondysmorphic subtypes in discovery and replication cohorts. *P* values indicate the probability that the yield of CSVs in nondysmorphic ASD is not lower than that of dysmorphic ASD (one-sided, t-test). c) Inheritance of polygenic risk for ASD in dysmorphic and nondysmorphic ASD subtypes in discovery and replication cohorts. Box plots depict the median and quartiles of pTDT deviation. Dots represent pTDT deviations of subjects. *P* values for each subgroup indicate the probability that the mean of the pTDT deviation distribution is not greater than zero (one-sided, t-test), as depicted by the dotted line. The finding of no significant over-transmission in dysmorphic ASD did not replicate in SSC, which might be due to lack of statistical power (i.e., at least 100 dysmorphic samples are needed to achieve 80% power if PRS explains 2.45% of phenotypic variance^11^) and/or ascertainment differences between the discovery and replication cohorts. The discovery cohort included data about major congenital anomalies in morphologic classification, whereas the replication cohort did not. While our discovery cohort was population-based, the Simons Simplex Collection excluded probands with medically significant perinatal diseases, severe neurological deficits, and certain genetic syndromes^27^. This likely decreased the proportion of probands with excess MPAs and birth defects, potentially leading to a lower burden of common ASD-associated variants^85^.

In both cohorts we also found a significant over-transmission of common ASD-associated SNPs in ADM-defined nondysmorphic ASD (*P*_discovery=_ 6.7×10^−3^ and *P*_replication=_ 6.3 ×10^−3^, one-sided Wilcoxon rank sum test, Figure 6c). In results similar to Weiner *et al*.^8^, we did not observe over-transmission in unaffected siblings in the replication cohort (*P=* 0.88, one-sided Wilcoxon rank sum test).

Individuals with ADM-defined dysmorphic ASD or with CSVs had a significantly lower IQ compared to ADM-defined nondysmorphic ASD or those without CSVs, respectively (Supplementary Figure 4a and b, Supplementary Tables 12, 14-16). Although there was no correlation between IQ and GRVS in the discovery cohort when the subtype classification was done by either gold standard dysmorphology examination (rho= -0.042, *P=* 0.64, Figure 5c) or using the ADM (rho= 0.12, *P=* 0.21, Supplementary Figure 4c), a significant negative correlation was found in the replication cohort (rho= -0.14, *P=* 4.4×10^−3^, Supplementary Figure 4c). We did not find significant correlations between IQ and PRS (Figure 5d and Supplementary >Figure 4d), or PRS and GRVS in either cohort (Supplementary Figure 5 and Supplementary Tables 12 and 16).

Differences in the correlation between GRVS and IQ between the cohorts might be attributable to differences in ascertainment. The discovery cohort was assembled using a population-based recruitment strategy, and the average IQ of the cohort is 105 similar to the population average of 100. In contrast, individuals with comorbid ID or low IQ are found in SSC^29^, consistent with the replication cohort having a significantly lower IQ compared to the discovery cohort (mean IQ_discovery=_ 105±23, mean IQ_replication=_ 82±27, *P=* 1.1 × 10^−21^, two-sided t-test). Inconsistent findings between ASD cohorts have also been observed when examining gender differences in IQ^30^, where findings from cohorts with specific selection criteria (e.g., simplex families) may not be generalizable to the ASD population.

Our data suggest that while both dysmorphic and nondysmorphic ASD demonstrate over-transmission of common ASD-associated variants, there is a significantly higher burden of rare variants in dysmorphic ASD than nondysmorphic ASD. GRVS methods may add further specificity to identifying clinically informative endophenotypes but exquisitely phenotyped cohorts will be required. While dysmorpholgy classification by expert clinical examination is not highly scalable, the use of automated tools for 2 and 3-dimensional imaging^31^ may make it feasible to perform high throughput dysmorphology classification. This will allow GRVSs to be more widely used, potentially in combination with one or more early clinical biomarkers.

## Online Methods Subjects and Methods

### Subject enrolment – Discovery Cohort

The cohort consists of children residing in the Canadian province of Newfoundland and Labrador, recruited from one of three developmental team assessment clinics between 2010 and 2018. Assessment through one of these clinics was required for a child with ASD to qualify for provincially funded home Applied Behavioural Analysis (ABA) therapy. Families were invited to participate after their child received an ASD diagnosis from the multidisciplinary team which was led by a developmental paediatrician. Probands met ASD criteria according to the Diagnostic and Statistical Manual of Mental Disorders (Fourth or Fifth Edition, Text Revision)^32-34^ and all diagnoses were confirmed by an Autism Diagnostic Observation Schedule^35^ assessment. Most probands also had an Autism Diagnostic Interview-Revised^36^ assessment consistent with ASD. Children were not excluded from the study based on syndromic features or the presence of a known syndrome. Parents or guardians of the children provided written informed consent. The study was approved by Newfoundland’s Health Research Ethics Boards (HREB# 2003.027) and SickKids Research Ethics Board (REB#0019980189).

### Subject enrolment – Replication Cohort

The replication cohort consisted of a subset of samples from the Simons Simplex Collection, including 442 affected probands with dysmorphology and WGS data along with their unaffected siblings (n= 355).

### Clinical Assessment and Morphological Examination – Discovery Cohort

Clinical assessments, morphological examinations and classification were performed as previously described^9,10^. In brief, the team reviewed the child’s family history and medical records, including radiology and electroencephalogram (EEGs). EEGs were ordered if there was a clinical suspicion of seizures. Other screens for birth defects were arranged based on standard physical examination of the proband, which included a cardiovascular examination (e.g. echocardiogram for a proband with a murmur consistent with a ventricular septal defect). A single experienced dysmorphologist (B.A.F.) performed a detailed morphological examination of the child and (if possible) parents documenting minor physical anomalies (MPAs), height, weight, head circumference and anthropometric measurements of the head, face, hands, and feet. As described by Miles, *et al*.^9^, each proband was assigned an MPA score; one point was given for each embryologically unrelated MPA or for each measurement greater than two standard deviations above or below the population mean and that was absent from the parents if they were available for examination. Each child was also assigned a major congenital anomaly (MCA) score (two points were given for each MCA), and a total morphology score (MPA + MCA scores). Using the total morphology score, we classified each child into essential (total morphology score 0-3), equivocal (total morphology score 4-5) or complex (total morphology score ≥6) groups. We used the final classification for comparing the yield of CSVs and for performing the rare and common variant analyses.

### Autism Dysmorphology Measure – Discovery and Replication Cohorts

Our replication cohort consisted of a subset of samples from the Simons Simplex Collection^27^. This subset of samples had already been categorized into two morphological groups (400 nondysmorphic and 42 dysmorphic cases) by multiple non-geneticist examiners using the Autism Dysmorphology Measure^28^. In brief, the Autism Dysmorphology Measure is a decision tree-based classifier that assigns cases into nondysmorphic and dysmorphic groups based the presence or absence of minor physical anomalies of 12 body areas. It was designed to be used by clinicians who do not have expert training in dysmorphology and the assessment is limited to the craniofacies, hands and feet of the child. The ADM decision tree was trained on expert-derived consensus classification of 222 ASD cases who had gold standard examinations of all body areas by clinical geneticists with expertise in dysmorphology^9,37^. The latter was the approach we used for the initial morphologic classification of our discovery cohort into essential, equivocal and complex groups^9^.

In contrast to the Autism Dysmorphology Measure, the morphological scores used to classify the discovery cohort factored in major congenital anomalies as well as MPAs, and MPAs were documented for the entire body including areas not assessed by the ADM (for example the thorax, arms, legs and skin). In order to align the type of morphologic data that was used to classify the discovery and replication cohorts, we reclassified the discovery cohort using the Autism Dysmorphology Measure, yielding 248 nondysmorphic and 77 dysmorphic cases. Of the 248 ADM-defined nondysmorphic cases, 18 cases were clearly dysmorphic upon further review by an experienced dysmorphologist (B.A.F). The Autism Dysmorphology Measure is reported to have an 82% sensitivity^28^, and the sensitivity for the discovery cohort is similar at 80%. Thus, we excluded the 18 individuals with a false negative dysmorphic ADM classification to make the discovery cohort as clean as possible. We also included only samples that were sequenced on Illumina platforms to be consistent with the replication cohort^27^.

Thus, the final number of ASD cases in the discovery cohort used for analysis was 276, of which 203 had nondysmorphic ASD and 73 had dysmorphic ASD according to ADM.

### Whole-genome sequencing and variant detection

We extracted DNA from whole blood or lymphoblast-derived cell lines and assessed the DNA quality with PicoGreen™ and gel electrophoresis. We sequenced 795 genomes (325 probands and 470 parents) with one of the following WGS technologies/sites as previously described^4^: Complete Genomics (Mountain View, CA, n= 33 probands, 64 parents), Illumina HiSeq2000 by The Centre for Applied Genomics (TCAG) (Toronto, ON, n= 24 probands, 48 parents), or Illumina HiSeq X by Macrogen (Seoul, South Korea, n= 182 probands, 250 parents) or TCAG (n= 86 probands, 108 parents). We used KING^38^ to confirm familial relationships and ADMIXTURE^39^ and EIGENSOFT^40^ to confirm ancestries (Supplementary Table 12).

Alignment and variant calling for genomes sequenced by Complete Genomics were conducted as previously described^41^. For samples sequenced on Illumina platforms, each WGS site aligned WGS reads to the human reference genome assembly hg19 (GRCh37) using Burrows-Wheeler Aligner v.0.7.12^42^ (TCAG) or Isaac v.2.0.13^43^ (Macrogen). For each genome, we performed local realignment and quality recalibration and detected SNVs and small indels using the Genome Analysis Toolkit (GATK) Haplotype Caller^44^ v.3.4.6 without genotype refinement. We detected CNVs using ERDS (estimation by read depth with single nucleotide variants)^45^ and CNVnator^46^ as previously described^47^. We detected SVs using Manta v.0.29.6^48^. When supported by the variant caller (i.e. GATK and Manta), trio-based joint variant calling was conducted for each family.

To identify uniparental isodisomies (isoUPDs), we calculated the ratio of the number of homozygous or hemizygous SNPs to the number of SNPs per chromosome, for each sample. Samples with a ratio greater than 0.55 had a putative isoUPD on the corresponding chromosome. We examined CNV and kinship data to rule out confounding factors (i.e., large CNVs or consanguinity). For each sample with a ratio greater than 0.55, we examined plots of B-allele frequency per chromosome; those with runs of homozygosity > 10Mb on one chromosome were considered to have a putative isoUPD^49^. We examined the inheritance of homozygous SNPs within the region of the putative isoUPD via visual inspection of BAM files and experimentally validated one of the SNPs to confirm the isoUPD and inheritance.

We systematically detected aneuploidies by calculating a ratio of the average read depth per chromosome to that for the entire sample. Ratios ≤0.5 and ≥1.5 were considered a loss or gain, respectively. For Complete Genomics data, we identified aneuploidies by looking for an excess of large CNVs for each chromosome per sample.

Tandem repeats were detected from samples with PCR-free DNA library preparation and sequenced on the Illumina HiSeq X platforms using ExpansionHunter Denovo^50^ with default parameters. We detected tandem repeat expansions in the discovery cohort using ExpansionHunterDenovo size cutoffs as previously described^51^. Sample quality control procedures were performed as previously described^51^.

### Variant Annotation

We annotated SNVs and indels with information on population allele frequency, variant impact predictors, and putative pathogenicity and disease association, using a custom pipeline based on ANNOVAR^52^ as previously described^4^. For non-genic regions, we annotated whether the variant overlapped reported ASD-associated non-coding regions^19-23^ (Supplementary Table 19). These included transcription start sites, fetal brain promoters and enhancers of LoF intolerant genes^20^, histone modification (H3K27ac) sites in fetal and adult brain^21^, splice sites, 3’- and 5’-untranslated regions (UTRs)^23^, binding sites predicted by DeepSEA^22^ to cause LoF, as well as conserved promoters of any genes, developmental delay-associated genes, and long non-coding RNA genes^19^. We tested three additional functional sites that have not been previously associated with ASD. These included boundaries of topologically associating domains^53^, CTCF binding sites^54^, and brain enhancers from Roadmap Epigenomics chromatin states (15-states chromHMM)^55^.

We annotated CNVs and SVs with a custom pipeline using RefSeq gene models, with repeat regions, gaps, centromeres, telomeres and segmental duplications relative to University of California at Santa Cruz genome assembly hg19. Similar to our non-genic annotations for SNVs, we annotated whether a CNV overlapped promoters of genes^19^, H3K27ac sites^19^, 3’UTR and 5’UTR^23^ (Supplementary Table 19). We retained CNVs overlapping such regions, but not exonic regions. We also annotated the frequency of each CNV and SV from among 3,107 parents in the MSSNG database^4^ (fifth version) and the putative pathogenicity and disease association [from Human and Mouse Phenotype Ontologies^56,57^ (HPO and MPO), ClinGen Genome Dosage Sensitivity Map^58^, Online Mendelian Inheritance in Man, and Database of genomic variation and phenotype in humans using ensemble resources (DECIPHER)^59^].

We annotated mitochondrial variants using Annovar-based custom scripts with annotations from MitoMaster (April 2019) and Ensembl v96.

### Detection of rare variants

We extracted high quality rare data for SNVs and indels after applying the following filters: 1) FILTER is PASS or varQuality is VQHIGH or PASS; 2) population allele frequencies < 1% in 1000 Genome Project^60^, NHLBI-ESP^61^, Exome Aggregation Consortium^62^, The Genome Aggregation Database^63^, and internal Complete Genomics control databases; 3) reference and alternative allele frequency > 95% and < 1%, respectively, based on allele frequencies of 2,573 parents in MSSNG (fourth version)^4^ to decrease batch and cross platform effects; and 4) allele frequency < 5% from 250 parents from this study aligned with Isaac to decrease alignment-specific artifacts. To further minimize cross platform and batch effects, we required heterozygous SNVs and indels to have an alternative allele fraction of 0.3-0.7 (inclusive) and homozygous/hemizygous SNVs and indels to have an alternative allele fraction >0.7 for variants from Complete Genomics. For Illumina variants, we also required heterozygous SNVs and indels to have a genotype quality score of at least 99 and 90, respectively, and homozygous SNVs and indels to have a genotype quality score of at least 25.

We retained CNVs >2kb that had <70% overlap with gaps, centromeres, telomeres, and segmental duplications. For CNVs from Illumina platforms, we defined stringent CNVs as those called by both ERDS and CNVnator (with 50% reciprocal overlap). We defined CNVs as rare if the allelic frequency was < 1% in parents from the MSSNG database^4^ and < 5% in parents of this cohort that were aligned with Isaac.

We retained as rare SVs, those with an allelic frequency of < 1% in parents analyzed with Manta from the MSSNG database and <5% in parents in this cohort that were aligned with Isaac. Pairs of entries with identical non-zero first numbers in the MATEID tag were retained as one inversion. Entries with identical MATEID values were retained as complex SVs. On average per sample, we detected ∼3.7 million SNPs, 36,514 rare single nucleotide variants (SNVs), 4,113 small insertions and deletions (indels), 13 rare copy number variants (CNVs), 390 rare structural variations (Supplementary Table 2).

### Detection of de novo variants

We determined *de novo* SNVs and indels from Complete Genomics data as previously described^41^. For Illumina WGS data, we also used DenovoGear^64^ (version 0.5.4) to detect *de novo* SNVs and indels. We extracted variants inconsistent with Mendelian inheritance (present in offspring but not in parents) with FILTER= PASS and defined rare, as above. To identify high confidence *de novo* SNVs, we applied the following quality filters: 1) pp_DNM score ≥ 0.9 from DenovoGear^64^; 2) overlap GATK^44^ calls with genotype quality scores ≥ 99 for heterozygous SNVs. We defined high confidence *de novo* indels as those called by DenovoGear and GATK with the same start site. We retained *de novo* SNVs and indels with a ratio of sequenced reads supporting the alternative call to the total number of reads at the position of 0.3-0.7, or > 0.7 for X- and Y-linked variants not in the pseudoautosomal regions in male subjects.

We defined putative *de novo* CNVs as rare stringent CNVs (see “Detection of rare variants”) that were inconsistent with Mendelian inheritance. For CNVs that did not have a conclusive inheritance pattern (i.e., CNV in child and parent were not the same size), we defined putative *de novo* CNV as those with a CNV length ratio between child and parent of > 2. For each putative *de novo* CNV from Illumina platforms, we calculated a read depth ratio of the CNV with the surrounding region in each family member, as previously described^47^. Ratios of 0.35-0.65 were considered heterozygous deletions, <0.35 as homozygous/hemizygous deletions, >= 1.4 as duplications and 0.9-1.1 as a normal copy number. Putative *de novo* CNVs were considered *de novo* if the copy number status based on ratios were inconsistent with Mendelian inheritance. For the 40 regions with ratios that did not meet the afore-mentioned criteria, we visualized the WGS reads to determine the inheritance status for samples sequenced by Illumina. To determine the inheritance status for samples sequenced by Complete Genomics, we examined the read depth coverage of the CNV relative to that of Complete Genomics controls^65^ and its flanking regions in each family member. On average per sample, we detected 73.4 *de novo* SNVs, 7.3 *de novo* indels, and 0.1 *de novo* CNVs (Supplementary Table 2)

### Validation of variants

We randomly selected a subset of all high quality exonic *de novo* SNVs, all *de novo* indels and all CSVs for validation in probands and available parents. We used Primer3^66^ to design primers to span at least 100 bp upstream and downstream of a putative variant, avoiding regions of known SNPs, repetitive elements, and segmental duplications. DNA from whole blood, if available, was used to amplify candidate regions by polymerase chain reaction and to assay with Sanger Sequencing. For CNVs, we validated all high confidence *de novo* exonic and all clinically significant CNVs in whole blood DNA (if available) of probands and available parents using TaqMan™ Copy Number Assay (Applied Biosystems), SYBR ^®^ Green qPCR (Thermofisher) or digital droplet PCR (BioRad). Experimental validation rates were 94.8%, 85.7%, and 87.5%, respectively, for *de novo* SNVs, indels, and CNVs (Supplementary Tables 3 and 4).

### Mitochondrial variant detection

For the samples sequenced by Illumina platforms, reads aligning to the mitochondrial genome were extracted and realigned to the revised Cambridge Reference Sequence (NC_012920) in b37 using BWA v0.7.8. Pileups were generated with samtools mpileup v1.1 requiring the program to include duplicate reads in the analysis and retaining all positions in the output. Custom scripts were developed to parse the mpileup output to determine the most frequently occurring non-reference base at each position in the mitochondrial genome. The heteroplasmic fractions were calculated and vcf files were generated. Fasta files with the most frequently occurring base at every position were also generated and used as input for the program HaploGrep v2.1.1 for haplogoup prediction. The vcf files were annotated using Annovar based custom scripts with annotations from MitoMaster (April 2019) and Ensembl v96.

For the samples that were sequenced by Complete Genomics, the mitochondrial variants called by the proprietary software were extracted. Fasta files were generated using custom scripts replacing mitochondrial reference bases with alternative bases at heteroplasmic sites and the files were used as input for the program HaploGrep v2.1.1 for haplogoup prediction. The vcf files were annotated using Annovar based custom scripts with annotations from MitoMaster (April 2019) and Ensembl v96.

Positions with heteroplasmic fraction less than 5% or greater than 95% and common in certain haplogroups (greater than 5%) were excluded from downstream analysis. All variants were manually reviewed, and a list of artefacts was compiled and excluded. To identify pathogenic mitochondrial variants, the following variants were considered: any MitoMaster pathogenic variants at 5-100% heteroplasmy, variants between 10-90% heteroplasmy, and variants between 5-100% heteroplasmy and seen <2% of the time in the individual’s haplogroup.

### Variant detection for replication cohort

For the replication cohort, CRAM files and sequence-level variants were downloaded from Globus (https://www.globus.org/). We detected CNVs using ERDS ^45^ and CNVnator^46^, as previously described^47^. Rare variants were filtered as described for the discovery cohort. We identified *de novo* SNVs and indels using DeNovoGear^64^. Allele frequencies from the Simons Simplex Collection were calculated and *de novo* variants with internal frequencies <1% were excluded. *De novo* SNVs and indels at poorly sequenced or highly variable sites were also excluded from further analysis. The remaining *de novo* variants were filtered as described for the discovery cohort with the exception of using a PP_DNM <0.95 threshold for *de novo* SNVs. Variants were annotated as described above for the discovery cohort.

### Variant prioritization and molecular diagnosis

To identify CSVs from the discovery cohort, we prioritized rare and *de novo* LoF and damaging (as predicted by at least five/seven predictors^23^) missense variants, and variants reported by ClinVar^67^ or the Human Gene Variant Database^68^. We also prioritized rare and *de novo* CNVs and SVs, including those overlapping syndromic regions in DECIPHER^59^ or ClinGen Genome Dosage Sensitivity Map^58^ databases. Genes affected by such variants were compared to ASD candidate genes^3,4,13,69,70^, candidate genes for neurodevelopmental disorders^69^, and genes implicated in neurodevelopmental or behavioural phenotypes according to HPO^57^ and MPO^56^. Additionally, we considered the mode of inheritance from the Online Mendelian Inheritance in Man and Clinical Genomics Database^70^, segregation and genotype-phenotype correlations. We classified the variants as pathogenic, likely pathogenic, variants of uncertain significance, likely benign, or benign, based on the American College of Medical Genetics and Genomics Guidelines^16,17^. Variants of unknown significance in known or candidate ASD genes with emerging evidence were further categorized into three ASD candidate variant categories (Supplementary Note and Supplementary Tables 5-7). Although applying quality filters for high confidence variants is important to minimize false positives for burden analysis, this can increase false negatives. Therefore, we also manually inspected WGS data when we identified CSVs that did not pass filtering criteria for high confidence variants.

Clinically significant variants classified as pathogenic or likely pathogenic or that were considered clinically relevant (i.e., prompting further clinical management) were reviewed by a medical geneticist in the context of the patient’s phenotype and family history. Relevant findings were reported back to families through a clinical geneticist. Differences in the yield of CSVs among the morphological groups were calculated using Fisher’s exact test.

To identify CSVs from the affected probands in the replication cohort, the afore-mentioned approach was applied to *de novo* LoF, damaging missense and CNVs. CSVs from the replication cohort were confirmed by manual inspection of WGS reads.

### Rare variant burden analysis in gene sets and noncoding regions

For the discovery cohort, we performed two ASD subtype comparisons for each rare variant burden analysis as follows: 1) comparing complex, equivocal and essential ASD using ordinal regression tests and 2) comparing complex and equivocal ASD (i.e. dysmorphic ASD) to essential ASD using logistic regression tests. The test was done by regressing an event (e.g., number of genes impacted by rare deletions per subject) capturing a particular genomic region (i.e., coding, gene sets, or noncoding regions) on the phenotype outcome (e.g., complex vs. essential ASD). The events tested in this study were the number of LoF, missense, and predicted deleterious variants for sequence-level variants and the number of genes or noncoding regions for CNVs. Tier 1 and 2 missense variants consist of all or only predicted damaging missense variants, respectively, as defined in Yuen, *et al*.^23^ The CNVs were grouped into two size bins, small CNVs (2-10kb) and large CNVs (10kb to 3Mb) due to greater proportion of these CNVs overlapping coding or noncoding regions, respectively. The number of genes impacted by other CNVs was based on their overlap with the coding regions of each gene. However, the number of genes impacted by small CNVs were based on genic overlap since there were not enough small coding CNVs for the gene set enrichment analysis. We compiled a list of 37 gene sets related to neuronal function, brain expression, mouse phenotypes from MPO, or human phenotypes from HPO that have been previously associated with ASD or used as negative control gene sets when comparing ASD to control groups (Supplementary Table 20) ^71-81^. For non-coding regions, we compiled a list of regions reported to be associated with ASD (Supplementary Table 19)^19-21^. We also included a score that predicts the impact of a variant on transcription factor binding as one of the non-coding regions tested^22^. Logistic regression and ordinal regression were applied for two subtypes and three subtypes comparison, respectively. Sex, genotyping platform, and three principal components from population stratification were included in the model as covariates to correct for any biases caused by sex difference, platforms, or ethnicity. Deviance test *P* value was calculated by comparing residuals from two regression models; one with just the covariates and another with all both covariates and target variable as previously described^82^. Global burden analysis was performed to compare the total number of LOF variants, missense variants, predicted deleterious variants for sequence-level variants, and genes impacted by CNVs. The coefficients reported were obtained from the model with the covariates. Multiple test correction for global burden tests was done using Benjamini Hochberg approach (BH-FDR). For the gene sets and noncoding regions burden test, total variant count (for SNVs and noncoding CNVs) or total gene count (for CNVs) was also included as one of the covariates to get rid of a global burden bias that might inflate the test *P* value. The coefficients, however, were calculated from the model with all the covariates except the total variant count or the total gene count for the actual magnitude of their impact. Permutation-based FDR correction (1000 permutations) corrected for the multiple comparison. Since different gene sets and non-coding regions consist of different number of genes or regions, we calculated the coefficients using z-scores for the number of features in each gene set/region to compare the coefficients across morphology-associated regions. When examining the burden of rare variants using logistic regression models, we used all probands from the discovery cohort (n=325). Since some probands did not have their parents sequenced, we used a subset of the discovery cohort (n= 235) when examining *de novo* variants.

### Genome-wide rare variant score

In addition to identifying relevant gene sets or regions that were differentially enriched among ASD morphologic subgroups, we developed a procedure to calculate a genome-wide rare variant score (GRVS) for each subject. This allowed the contribution of different variant types towards phenotype severity to be assessed together. The procedure involved two main steps: i) identification of relevant, differentially enriched gene sets or noncoding regions for each variant type along with an estimation of their effect sizes in the discovery cohort, and ii) calculation of the score for each subject in the target cohorts.

To estimate the effect sizes in the discovery cohort, we first fitted a logistic regression model by regressing platform, sex and first three principal components from population stratification on the dysmorphology classification (nondysmorphic= 0 and dysmorphic= 1, or essential= 0, equivocal= 1, complex= 2). We then used the regression coefficients of these covariates and the intercept in the second logistic regression model, where a feature representing a particular gene set or region was tested. Therefore, regression coefficients of all the gene sets and regions were corrected for those possible biases from the covariates equally. The two models can be notated as below

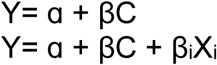

where Y is the outcome variable of dysmorphology classification, α is an intercept, β is a regression coefficients of covariates, C is a vector of covariates, β_i_ is the regression coefficient of a morphology-associated region, i, and X_i_ is the number of features found in a morphology-associated region. A feature is defined as the number of rare or *de novo* SNVs or indels or the number of genes or noncoding regions impacted by rare CNVs. For rare variants, we used all probands in the discovery cohort. Since some probands did not have their parents sequenced, we used a subset of the discovery cohort when examining *de novo* variants. To determine the optimal *P* value threshold to identify significant gene sets, we calculated the Nagelkerke’s R^2^ at different *P* value thresholds (*P* < 0.001, 0.005, 0.01, 0.05, 0.1. 0.5, and 1) using the discovery cohort and 10-fold cross-validation strategy. The optimal *P* value threshold was at P<0.1 (Supplementary Figure 3). To minimize the redundancy in significant gene sets and noncoding regions, we retained the most significant gene sets and noncoding regions with a Jaccard index < 0.75. We used the regression coefficients (β_i_) of significant gene sets or noncoding regions (*P* < 0.1) as a weight for the number of variants in those gene sets or regions in the GRVS calculation.

For each individual, the GRVS was calculated using the formula below

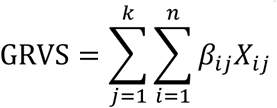

where n is the number of significant (*P* < 0.1) gene sets or regions for a particular variant type, j, k is the number of variant types (e.g., de novo missense variants), β_i_ is a regression coefficient of a significant gene set or region, i, and X_i_ is the number of variants (for SNVs and indels) or number of genes or regions (for CNVs) that are found in the significant gene set or region in the sample.

To examine the GRVS in the discovery cohort, we used a 10-fold cross validation strategy to avoid over-fitting. Using this strategy, the discovery cohort was randomly divided into 10 equally sized subsamples (stratified by subtypes). We calculated the GRVS of each sample in each subset using the effect sizes determined in the remaining nine subsets. To minimize stochasticity in the GRVS calculation, we repeated this procedure 30 times and the average GRVS and average number of variants for each sample were used for subsequent subtypes comparisons (Supplementary Figure 1a). For the replication cohort, we calculated GRVSs using significant gene sets and effect sizes derived from the discovery cohort (Supplementary Figure 1b). GRVS can be calculated for probands regardless of whether their parents have been sequenced. However, there would be a systematic difference in GRVSs in the discovery cohort if all probands were used because those whose parents have been sequenced includes scores from *de novo* variants, whereas probands whose parents have not been sequenced do not have scores from *de novo* variants. To ensure that the same variant types (including *de novo* variants) were included in each score for probands in the discovery cohort, GRVS was calculated only for probands whose parents had also both been sequenced. GRVSs were standardized within each cohort and subtyping method. We tested whether GRVS is higher in dysmorphic ASD compared to nondysmorphic ASD using a one-sided Wilcoxon’s Signed Ranked Test.

We used our ADM-reclassified cohort as the discovery cohort for several reasons: 1) In contrast to the MPAs (dysmorphology data) from SSC which were identified by multiple non-geneticist examiners, MPAs in the discovery cohort were documented by a single dysmorphologist with over 20 years of clinical experience (B.A.F.). MPA’s for children in the discovery cohort were then put through the ADM algorithim and the cases were classified as ADM-dysmorphic or ADM-nondysmorphic. This strategy allowed us to use very uniformly collected phenotypic data to derive the morphology-associated regions and effect sizes for GRVS calculation. 2) Our discovery cohort also contains more dysmorphic probands than SSC, which gives more power to identify morphology-associated regions (enriched in dysmorphic ASD). 3) Lastly, the discovery cohort was assembled using a population-based recruitment strategy so that the morphology-associated regions identified come from a patient collection representative of ASD as it exists at the level of primary care providers. In contrast there are ascertainment biases in SSC (e.g., simplex families and exclusion of severely affected/ syndromic probands) which might limit the generalizability of effect sizes and morphology-associated regions in a population-based cohort^30^.

We calculated a score for CSVs using the GRVS formula if the CSV was identified in a proband with two sequenced parents, and if the variant occurred in or overlapped one of the morphology-associated gene sets or noncoding regions so that an effect size was available for that variant. 46 CSVs were identified in 46 probands and17 of these met the above criteria allowing us to calculate a score for the variant. Of the remaining 29 CSVs, 15 were identified in probands where sequencing data was not available from both parents and 14 variants did not overlap a morphology-associated region.

### Common variant and PRS analysis

We examined the contribution of common SNPs among ASD subtypes. We calculated the PRS for each sample by deriving ASD summary statistics from a population-based genome-wide association study (GWAS) of 13,076 cases and 22,664 controls from the iPSYCH project ^11^. We calculated the PRS for BMI, which was a negative control due to its lack of association with ASD^24^, using BMI summary statistics from a population-based GWAS of 322,154 individuals of European descent from the GIANT Consortium^83^. We preprocessed the GWAS summary tables to fix the effect allele mismatch (swapped A1 and A2 alleles and converted its odds ratio) and to remove ambiguous SNPs (i.e., SNPs with A to T and C to G variations) and multi-allelic SNPs.

We conducted joint genotyping of BMI- and ASD-associated SNPs only on samples sequenced on Illumina platforms (200 probands and 400 parents). We could not re-genotype Complete Genomics data, so the samples were excluded from further analysis. We retained SNPs with a minor allele frequency > 0.05 and genotyping rate > 90%, of which 349,682 SNPs and 428,364 SNPs intersected with iPSYCH-ASD and GIANT-BMI SNPs passing suggested a p-value threshold (P value < 0.1 for ASD and P value < 0.2 for BMI) by Weiner et al.^2^, respectively. We then calculated PRSs using PRSice^84^ (parameters used: clump-kb 250, clump-p 1.000000, clump-r2 0.100000, info-base 0.9) using a p-value threshold of 0.1 for iPSYCH and 0.2 for GIANT BMI, as suggested by Weiner *et al*.^8^. After clumping, only 18,549 SNPs and 38,245 SNPs remained for PRS calculation for ASD and BMI, respectively. Using standard methods as previously described^11^, we calculated PRS for ASD for the SSC replication cohort using 26,067 SNPs with *P* value < 0.1 after the clumping step. The PRSs in both cohorts were standardized (with a mean of zero and standard deviation of one). We used the pTDT method^8^ and one-sided t-test to examine the over-transmission of common variants associated with ASD susceptibility among subtypes. Probands were used in the analysis if the probands were of European ancestry and if sequencing data was available from both parents.

## Data Availability

Access to FASTQ data for samples in the discovery cohort that were consented for MSSNG can be obtained by completing the data access agreement: https://research.mss.ng. Access to FASTQ data for samples in the discovery cohort not consented for MSSNG, as well as VCF files for sequence-level variants for all samples in the discovery cohort are available at European Genome-Phenome Archive (pending EGA link and accession number. Submission in process.). Access to data for the replication cohort can be obtained by completing data access agreement (https://www.sfari.org/resource/sfari-base), as was done for this study.

https://www.sfari.org/resource/sfari-base

## Acknowledgements

We thank the families for participation and The Centre for Applied Genomics for their analytical and technical support. We thank Lisa Strug, Andrew Paterson, and Delnaz Roshandel for analytical assistance. This work was funded by Autism Speaks, Autism Speaks Canada, the University of Toronto McLaughlin Centre, the Canada Foundation for Innovation, the Canadian Institutes of Health Research (CIHR), Genome Canada/Ontario Genomics Institute, the Government of Ontario, Brain Canada, Ontario Brain Institute Province of Ontario Neurodevelopmental Disorders (POND), and The Hospital for Sick Children Foundation. A.J.S.C. was supported throughout this research by Ontario Graduate Scholarship from the Government of Ontario, Restracomp Research Fellowship from The Hospital of Sick Children, and Autism Research Training Award and Frederick Banting and Charles Best Scholarship from CIHR. S.W.S holds the Northbridge Chair in Paediatric Research at the Hospital for Sick Children.

## Author Contributions

A.J.S.C., R.K.C.Y., S.W.S., and B.A.F. conceived and designed experiments. B.A.F, C.N., T.N.T., and J.H.M. managed, recruited, diagnosed and examined participants. E.A. and R.P. helped with interpreting phenotype data. Z.W., B. Thiruvahindrapuram, B.Trost, T.N., G.P., W.S., and J.M. processed whole-genome sequencing data. A.J.S.C., W.E., R.K.C., D.M., D.R. and M.Z. conducted or interpreted different components of whole genome sequencing analyses. A.J.S.C., M.S.R., D.J.S., N.S. and K.T. performed variant interpretation. A.J.S.C. and S.L. performed experiments for variant characterization and validation. A.J.S.C., B.A.F., S.W.S., and R.K.C.Y. conceived and coordinated the project and wrote the manuscript.

## Code availability

Code used in this manuscript is available at GitHub (http://github.com/naibank/GRVS_ASD).

