## Supplementary Information for "Genome-wide rare variant score associates with morphological subtypes of autism spectrum disorder"

### Supplementary Note

#### ASD candidate variants:

We also identified 29 variants of unknown significance of interest in 26 probands that fell into three categories: 1) variants in known ASD/neurodevelopmental genes, but with unknown impact on gene function or disease, 2) variants in ASD/neurodevelopmental candidate genes with emerging evidence, or 3) tandem repeat expansions in previously reported ASD candidate loci[1]. Additional information regarding ASD candidate variants in category 1 are described below:

<u>Paternally inherited loss-of-function (LoF) variant in *CIC* in subject 3-0328-000</u>

*Cic*$^{+/-}$ mice exhibit mild hyperactivity compared to wild-type mice. Mice with conditional knockouts of *Cic* in forebrain show memory deficits, hyperactivity, altered cortical thickness, defects in neuronal maturation and maintenance, and altered dendritic branching. Conditional knockouts of *Cic* in hypothalamus and amygdala result in abnormal social interaction[2]. Five unrelated individuals with *de novo* LoF variants in *CIC* share similar clinical features, including intellectual disability, developmental delay, ASD, attention deficit and hyperactivity disorder, seizures, and brain abnormalities[2]. *De novo* LoF variants reported in Lu *et al*.[2] impact both *CIC* isoforms, whereas the paternally inherited LoF in our subject 3-0328-000 only affects the short isoform (*CIC-S*). The impact of this variant on *CIC* is unknown and there are no other reports of cases with only *CIC-S* impacted. However, the short isoform is expressed in mouse brain[3], The proband has complex ASD and intellectual disability; his phenotype is further described in Supplementary Table 7.

<u>Paternal uniparental isodisomy involving homozygous missense variant in *FAT4* in subject 3-0095-000</u>

A missense variant in *FAT4*, predicted to be damaging, was identified to be homozygous as a result of uniparental (paternal) isodisomy of chromosome 4. Homozygous LoF and missense variants of *FAT4* are associated with Van Maldergen Syndrome, which is characterized by intellectual disability, partially penetrant periventricular neuronal heterotopia, and craniofacial, skeletal, auditory and renal malformations[4]. He also has a pathogenic *de novo* LoF variant in *WAC*: a gene associated with Desanto-Shinawi syndrome, of which ASD is a main feature[5]. Given the phenotypes associated with these two syndromes, we suggest that the *de novo WAC* variant is a main contributor to his ASD phenotype, although we cannot rule out the possible additional impact of the homozygous missense *FAT4* variant. The proband has complex ASD; his phenotype is further described in Supplementary Table 7.

<u>Paternally inherited “templated sequence insertion” involving *PHF21A* and *GLIS2* in subject 3-0439-000</u>

*PHF21A* is within the chr11p11.2 deleted region associated with Potocki-Shaffer syndrome[6]. *De novo* LoF variants of this gene are associated with intellectual disability and craniofacial abnormalities, with ASD reported in 3 of the 10 reported cases[6-8]. Templated sequence insertions (TSIs) are characterized by reverse transcription of an RNA intermediate, LINE-1-based insertion, target site duplication, cryptic polyadenylation signal, and polyadenylation[9]. In subject 3-0439-000, the last intron and exon of *GLIS2* are inserted in an inverted manner into *PHF21A*, along with a polyadenylation sequence and microduplication of 17bp (Supplementary Figure 6). The impact of this TSI on *PHF21A* expression and function is unknown. The proband has equivocal ASD (see Supplementary Table 7).

<u>Inherited "templated sequence insertion" involving *FGFR2* and *NPM1* in subjects 3-0209-000 and 3-0728-000</u>

*FGFR2* is associated with several mutation-specific disorders (OMIM: 176943). Activating *FGRF2* variants are associated with several distinct craniosynostosis syndromes, some of which are associated with neurodevelopmental abnormalities. Subject 3-0209-000 has a paternally inherited TSI of *NPM1* cDNA into *FGFR2*. The insertion is inverted and is followed by a polyadenylation insertion and a microduplication. We found the same insertion as a maternally inherited TSI in subject 3-0728-000 (Supplementary Figure 7). It is unknown whether this variant affects the coding sequence of *FGFR2*, and whether this variant will be associated with a known disorder or a different disorder. Both probands have high functioning forms of ASD and neither has craniosynostosis. Subject 3-0209-000 has essential ASD. 3-0728-000 has complex ASD, attention deficit disorder and an anxiety disorder. (See Supplementary Table 7 for additional phenotypic information on the probands).

A

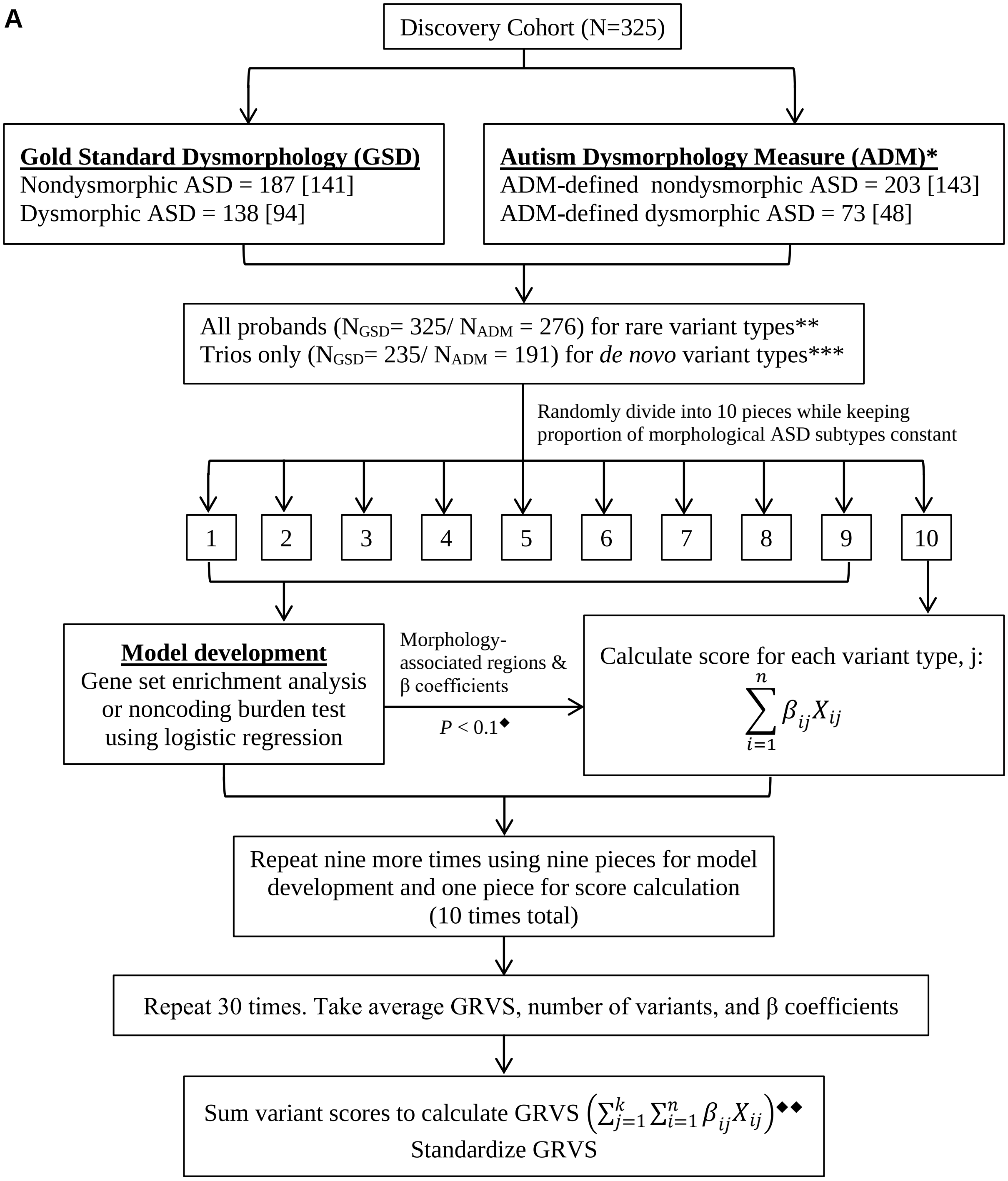

Discovery Cohort (N=325)
Gold Standard Dysmorphology (GSD)
Nondysmorphic ASD = 187 [141]
Dysmorphic ASD = 138 [94]
Autism Dysmorphology Measure (ADM)*
ADM-defined nondysmorphic ASD = 203 [143]
ADM-defined dysmorphic ASD = 73 [48]
All probands ($N_{GSD}$= 325/ $N_{ADM}$ = 276) for rare variant types**
Trios only ($N_{GSD}$= 235/ $N_{ADM}$ = 191) for *de novo* variant types***
Randomly divide into 10 pieces while keeping proportion of morphological ASD subtypes constant
1
2
3
4
5
6
7
8
9
10
Model development
Gene set enrichment analysis or noncoding burden test using logistic regression
Morphology-associated regions & β coefficients
$P < 0.1$◆
Calculate score for each variant type, j:
$\sum_{i=1}^{n} \beta_{ij} X_{ij}$
Repeat nine more times using nine pieces for model development and one piece for score calculation (10 times total)
Repeat 30 times. Take average GRVS, number of variants, and β coefficients
Sum variant scores to calculate GRVS $\left(\sum_{j=1}^{k} \sum_{i=1}^{n} \beta_{ij} X_{ij}\right)$◆◆
Standardize GRVS

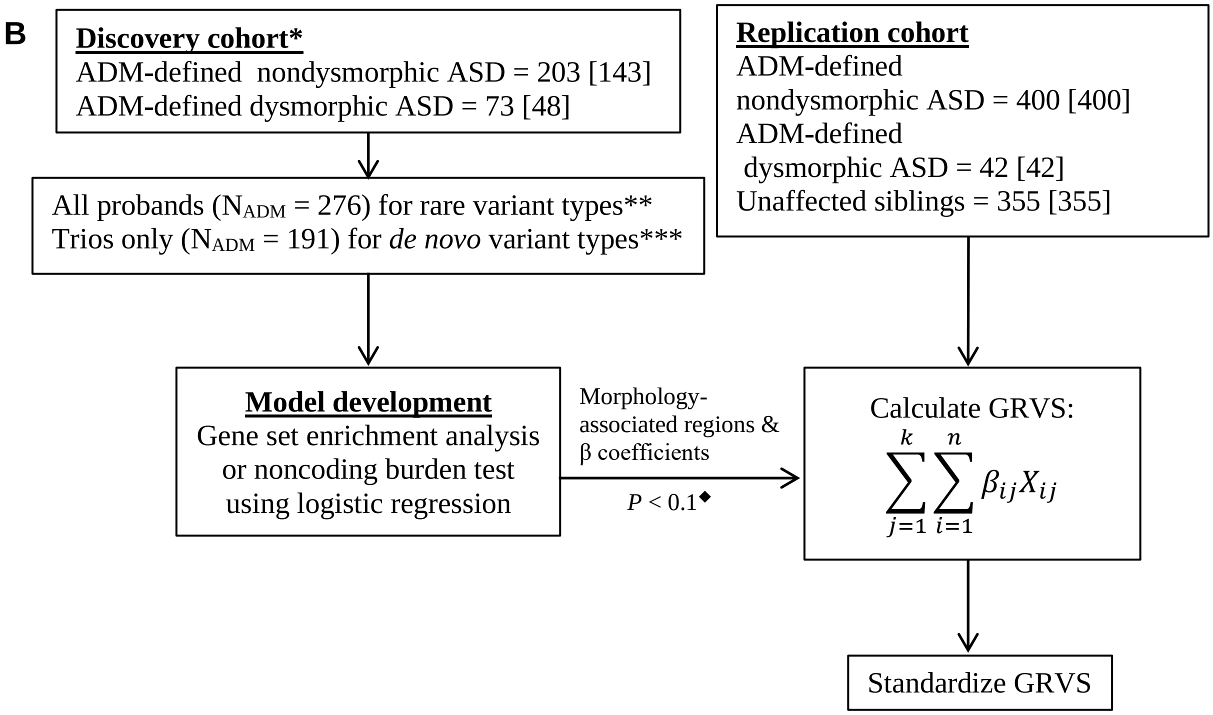


**Supplementary Figure 1: Flowchart for GRVS calculation for discovery and replication cohorts.** Square brackets indicate the number of trios in each subtype.

*Discovery cohort classified using ADM does not include false negative samples (i.e. complex ASD cases classified as ADM-defined nondysmorphic ASD), and only includes cases sequenced by Illumina.

**rare variant types that were analysed consisted of coding deletions >10kb, coding deletions ≤ 10kb, coding duplications >10kb, coding duplications ≤ 10kb, predicted loss-of-function variants, missense variants, predicted damaging missense variants, noncoding deletions >10kb, noncoding deletions ≤ 10kb, noncoding duplications >10kb, noncoding duplications ≤ 10kb, and noncoding SNVs and indels.

****de novo* variant types that were analysed consisted of predicted loss-of-function variants, missense variants, predicted damaging missense variants, and noncoding SNVs and indels.

□Nagelkerke's $R^2$ was calculated at different *P* value thresholds (*P* < 1, 0.5, 0.1, 0.01, 0.005, and 0.001) to determine the optimal *P* value threshold.

□□GRVS was calculated only probands with both parents sequenced because they had variant scores for both rare and *de novo* variants.

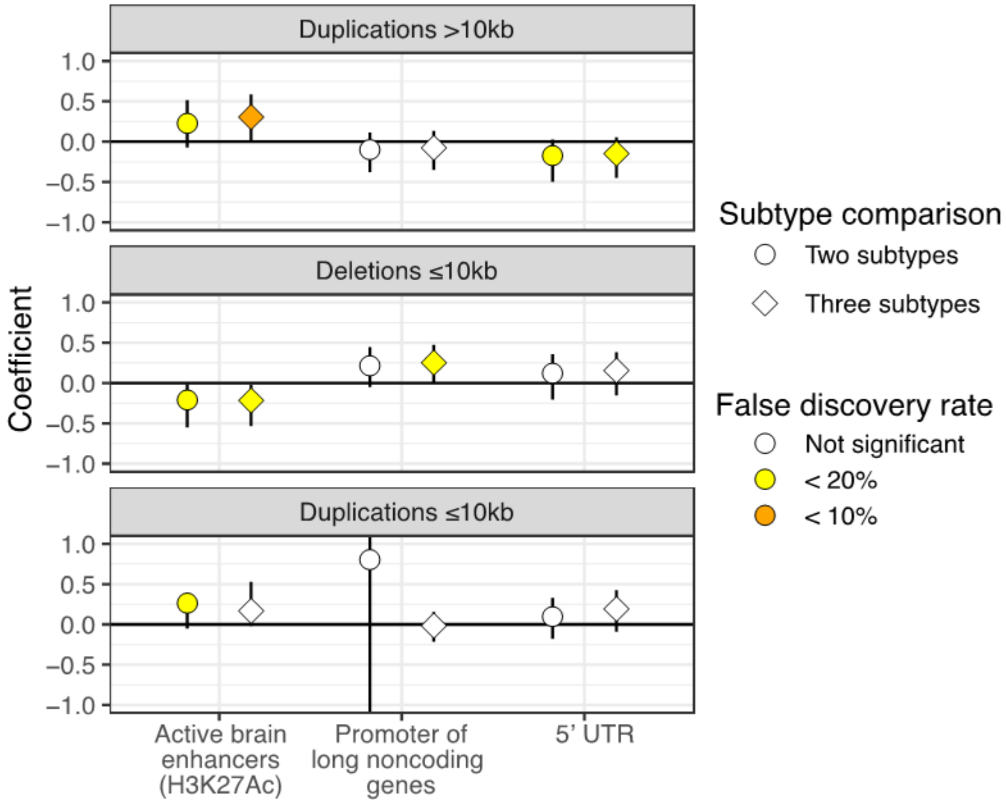


**Supplementary figure 2: Noncoding regions for which rare variants are significantly more prevalent in some subtypes of ASD.**

Events and coefficients are as described in Figure 2. We show only noncoding regions for which duplications >10kb (top panel), deletions ≤10kb (middle panel), and duplication ≤10kb (bottom panel) are significantly more prevalent in different subtypes of ASD. Symbol shapes indicate the subtype comparisons that were conducted for each combination of gene set and variant type. Two subtype comparison = nondysmorphic vs. dysmorphic ASD. Three subtype comparison = essential vs. equivocal vs. complex ASD. Coloured shapes indicate significant signals after multiple test correction by permutation-based FDR. Error bars indicate 95% confidence intervals.

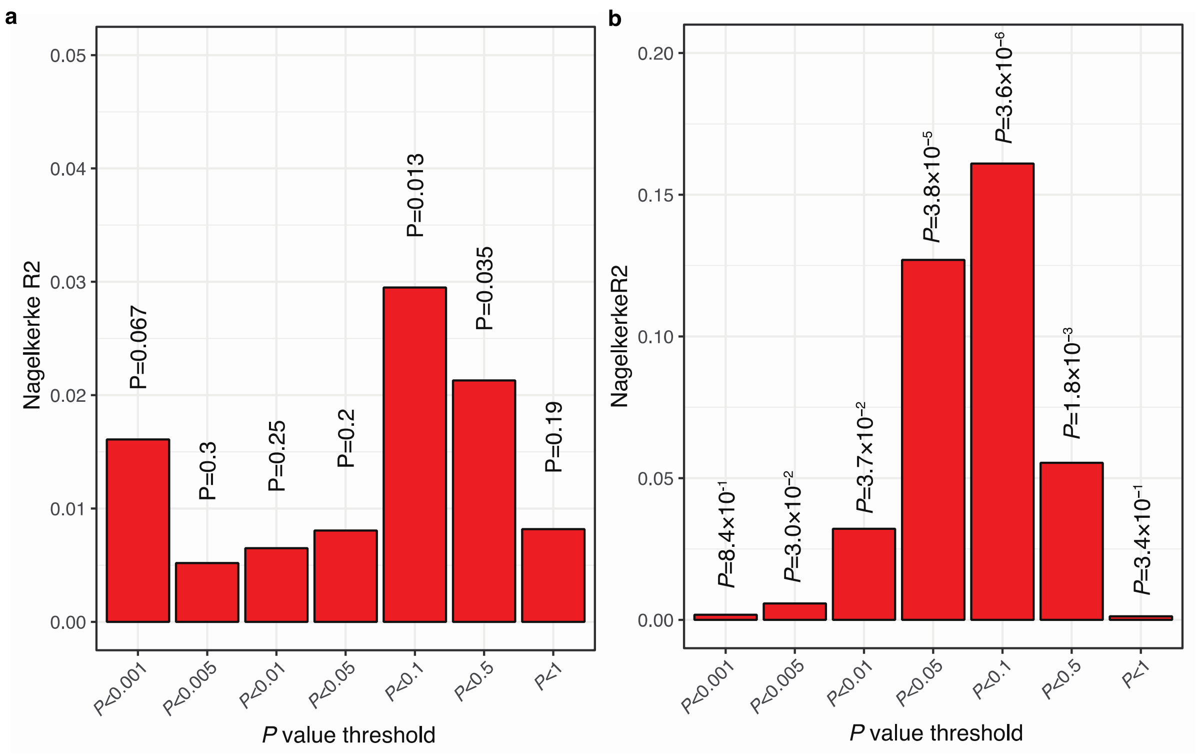


**Supplementary Figure 3: Nagelkerke's $R^2$ to determine optimal *P* value threshold for GRVS.** Shown is the distribution of Nagelkerke $R^2$ of GRVS in the discovery cohort using 10 × 30-fold cross validation on a) gold standard dysmorephology examinations, or b) Autism Dysmorphology Measure (ADM) at different *P* value thresholds, which were used to identify morphology-associated gene sets and noncoding regions for GRVS calculation. Based on both methods, the optimal *P* value threshold is *P*<0.1.

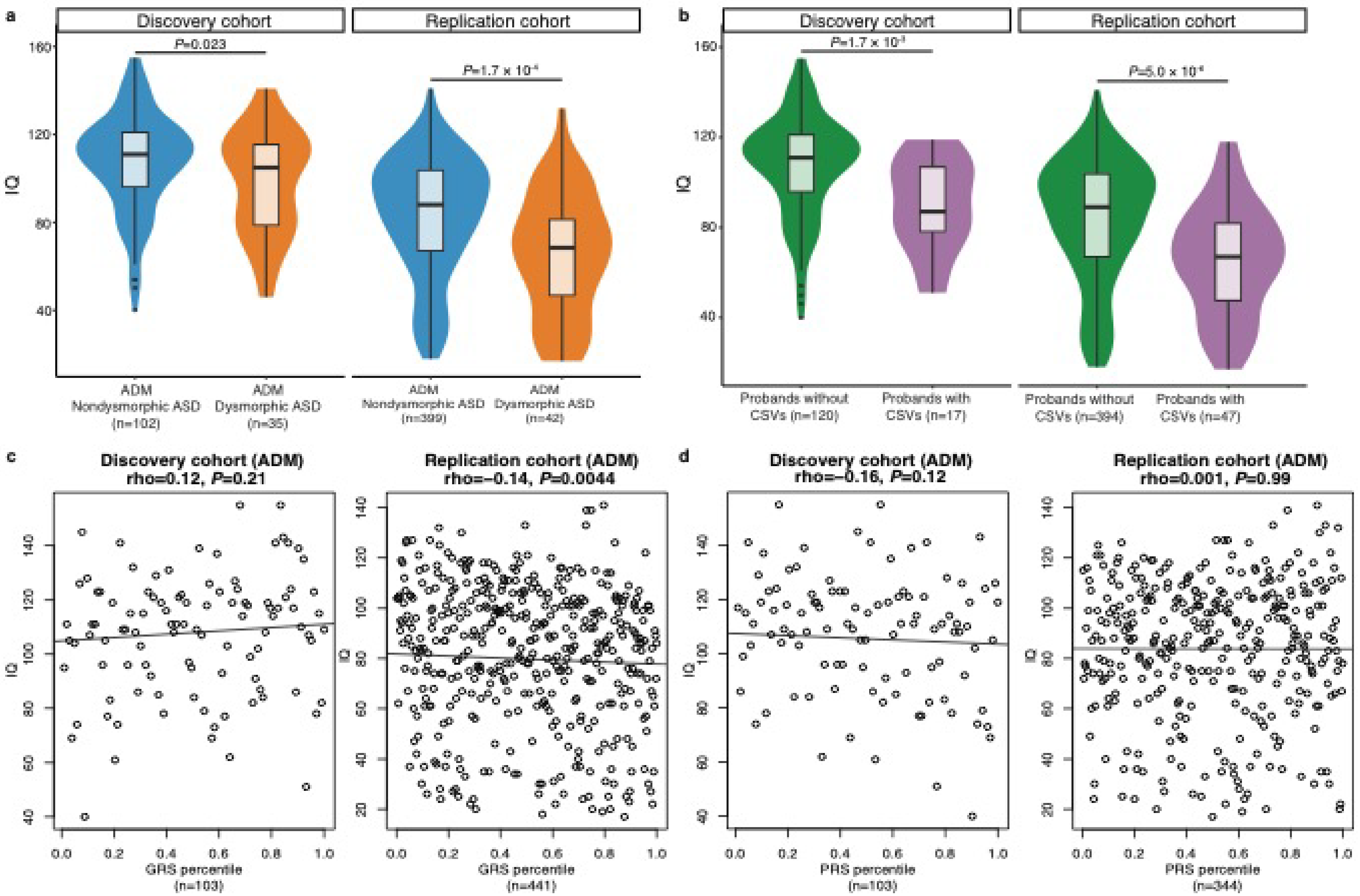


**Supplementary Figure 4: Relationship between IQ, genetic variants and morphological ASD subtypes classified by the Autism Dysmorphology Measure (ADM).** The left panels show results for the discovery cohort that was classified using the Autism Dysmorphology Measure, while the right panels show results for the replication cohort**.** IQ comparison between **a**) ASD subtypes classified by ADM, or **b**) probands with or without clinically significant variants (CSVs). Violin plots show the distribution of the samples' IQ; box plots contained within show the median and quartiles of IQ for each subtype. *P* values denote the probability that the mean IQ of ADM-defined nondysmorphic ASD or probands without CSVs is not greater than ADM-defined dysmorphic ASD or probands with CSVs, respectively (one-sided, t-test). Correlation between IQ and **c**) GRVS or **d**) PRS percentiles. Each dot represents the PRS and GRVS percentile for a sample in the discovery cohort or replication cohort. The linear regression line indicates the linear correlation between IQ and GRVS or PRS percentiles. Correlation coefficient is quantified by Spearman's rho correlation. *P* values indicate the probability that the correlation is occurred due to chance.

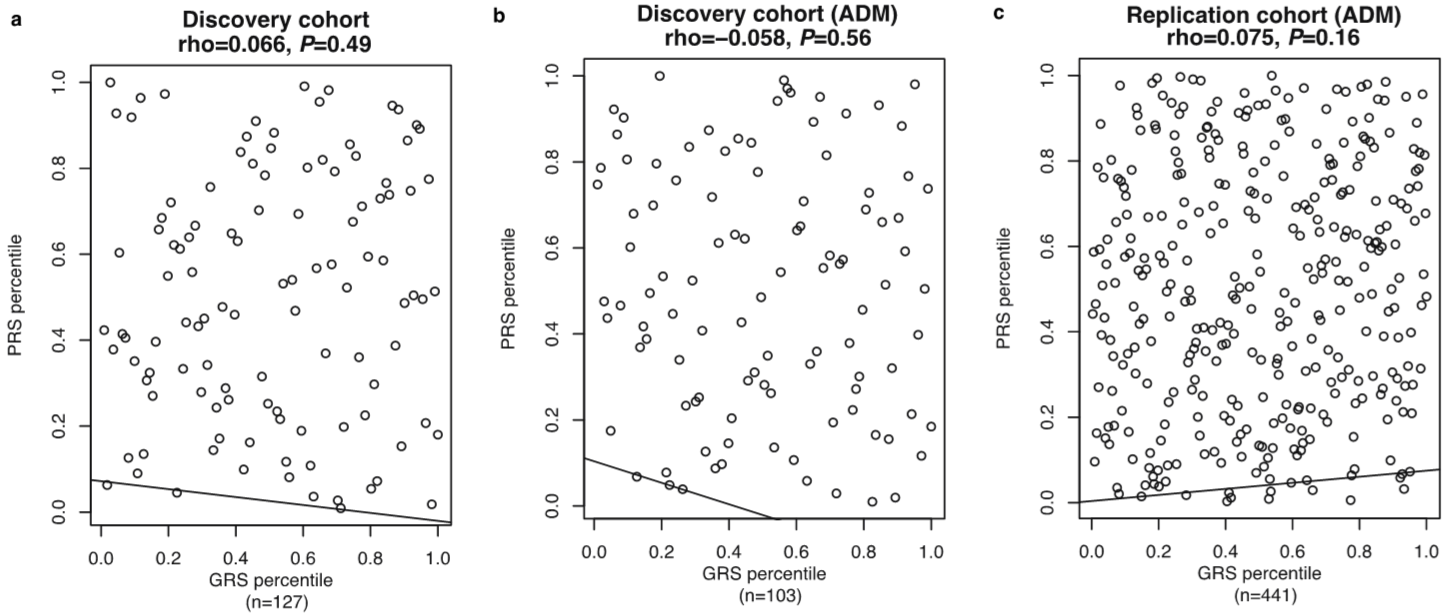


**Supplementary Figure 5: Correlation between GRVS and PRS.** Each dot represents the PRS and GRVS percentile for a sample in the discovery cohort using a) gold standard dysmorphology examinations, or b) the Autism Dysmorphology Measure (ADM), or c) in the replication cohort. The linear regression line indicates the linear correlation between GRVS and PRS percentiles. Correlation coefficient is quantified by Spearman’s rho correlation. *P* values indicate the probability that the correlation is occurred due to chance.

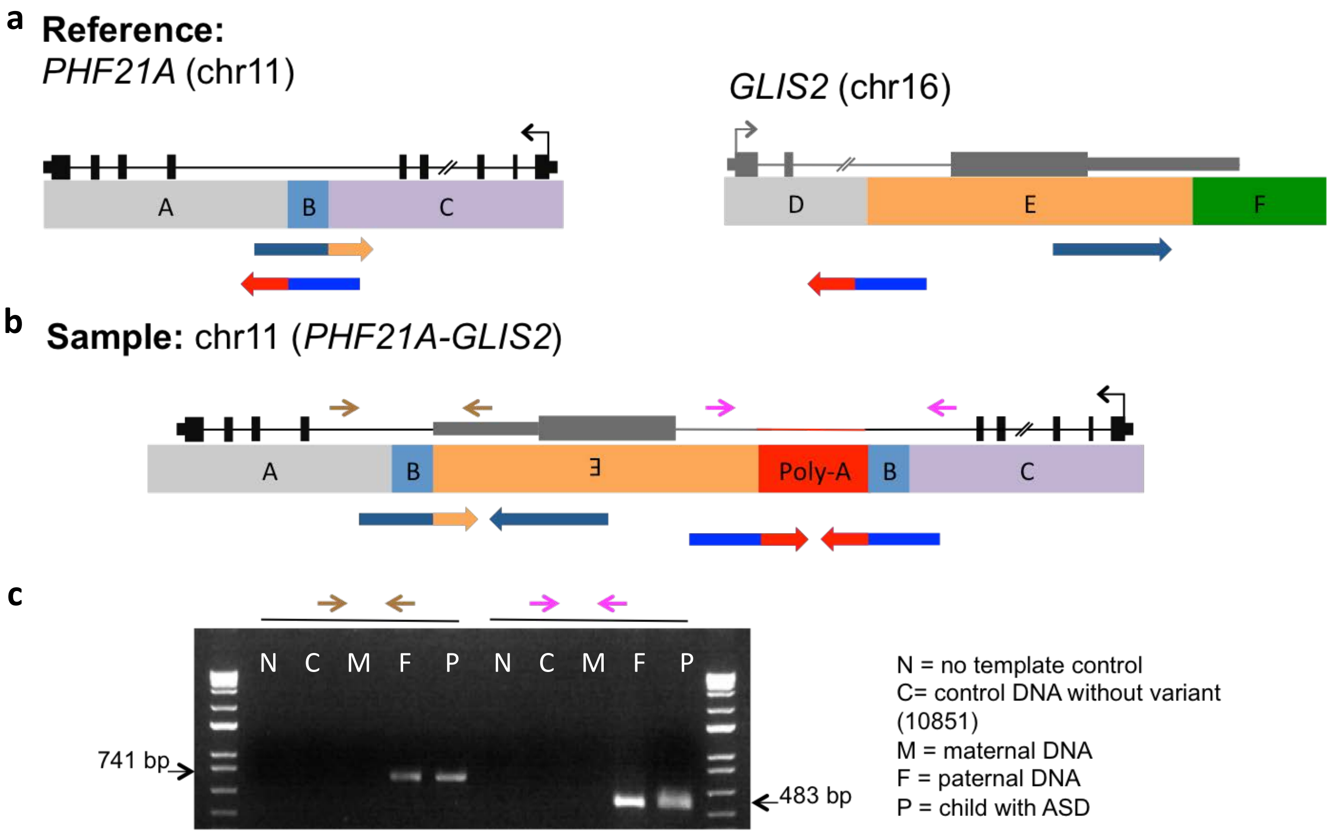


**Supplementary Figure 6: Illustration of paternally inherited templated sequence insertion of last intron and exon of *GLIS2* into intron 14 of *PHF21A* in subject 3-0439-000.**

**a**) Alignment of WGS reads (blue arrows) to *PHF21A* and *GLIS2* reference sequence (black and grey genes, respectively, and blocks A-F). Mate pairs are depicted by the same hue of blue. Split reads are depicted by the red and orange-coloured section on the blue reads that align to poly-A and section E, respectively. **b**) In the sample's genomic sequence, section E was duplicated and inserted into intron 1 of *PHF21A* along with a non-reference poly-A insertion (red block and line) and microduplication of section B. As a result, most of the last intron and exon of *GLIS2* (grey) were inserted into *PHF21A* (black genes). The sample's genes and genomic sequence is shown through black, grey and/or red lines and boxes, blocks A-C, E, and a poly-A block. Brown and magenta arrows depict the location of primers for PCR validation. **c**) PCR validation of variant allele in family 3-0439 at 5' and 3' end (brown and magenta arrows, respectively).

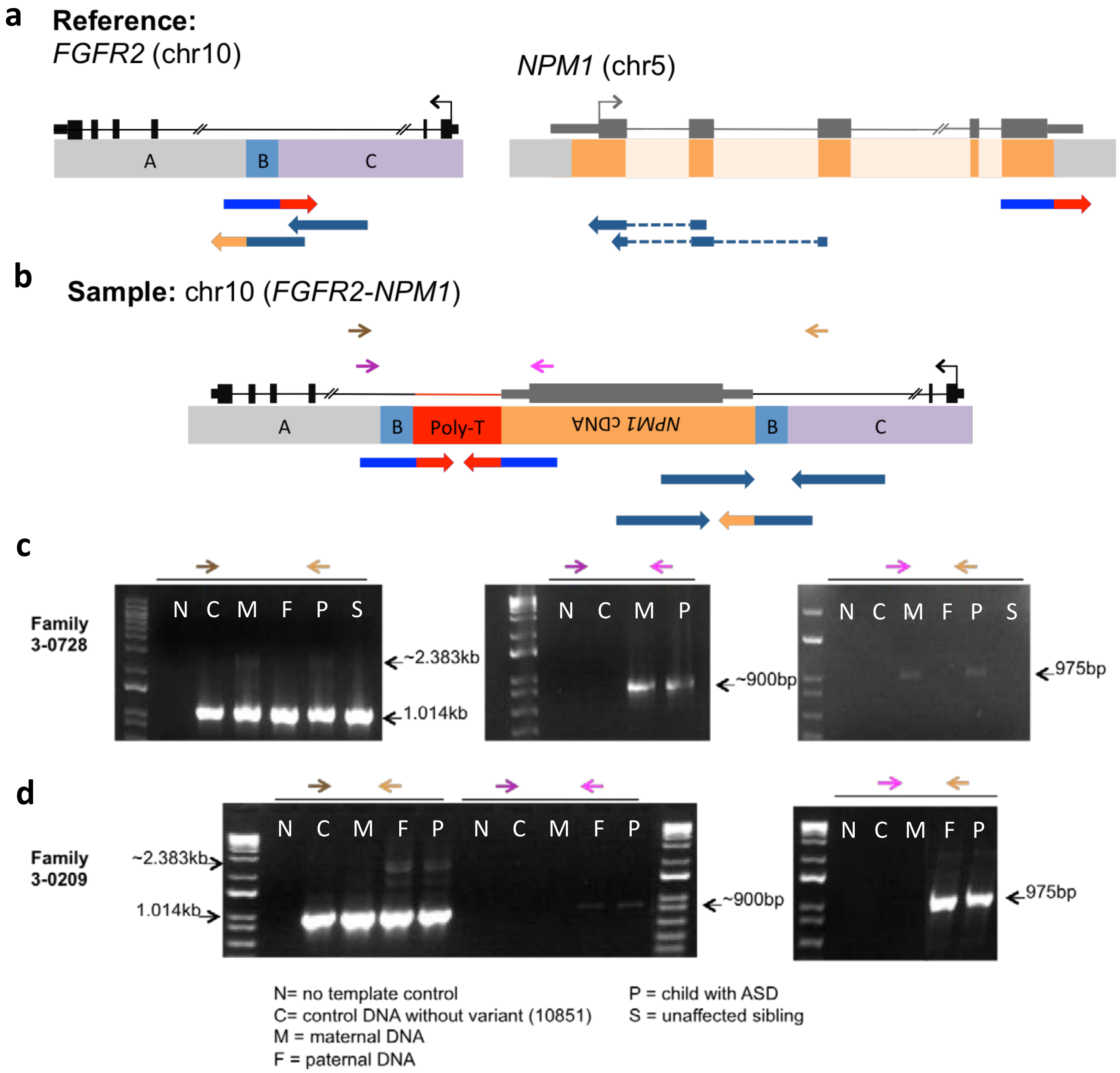


**Supplementary Figure 7: Illustration of insertion of *NPM1* cDNA into *FGFR2* in two subjects.**
**a**) Alignment of WGS reads (blue arrows) to *FGFR2* and *NPM1* reference sequence (black and grey genes, respectively). Mate pairs are depicted by the same hue of blue. Split reads are depicted by the red and orange-coloured section on the blue reads that align to poly-A and *NPM1* cDNA, respectively. Dotted lines indicate that the WGS read perfectly aligned to *NPM1* exons. **b**) In the sample's genomic sequence, most of *NPM1* cDNA (grey) was inserted into *FGFR2* intron (black) in an inverted manner along with a non-reference poly-A insertion (red block and line) and microduplication of section B. The sample's genes and genomic sequence is shown with black, grey, and red lines and boxes, blocks A-C, *NMP1* cDNA, and a poly-A block. Brown and magenta arrows depict the location of primers for

PCR validation. PCR validation of variant allele in families **c**) 3-0728 and d) 3-0209. Brown primer pairs were used to amplify DNA of each sample. Purple and magenta primer pairs were used to conduct nested PCR on the 5’ end of PCR products of brown primer pairs. Magenta and tan primer pairs were used to conduct nested PCR the 3’ end of PCR products of brown primer pairs.
